# A Novel Penalized Inverse-Variance Weighted Estimator for Mendelian Randomization with Applications to COVID-19 Outcomes

**DOI:** 10.1101/2021.09.25.21264115

**Authors:** Siqi Xu, Peng Wang, Wing Kam Fung, Zhonghua Liu

## Abstract

Mendelian randomization (MR) utilizes genetic variants as instrumental variables (IVs) to estimate the causal effect of an exposure variable on an outcome of interest even in the presence of unmeasured confounders. However, the popular inverse-variance weighted (IVW) estimator could be biased in the presence of weak IVs, a common challenge in MR studies. In this article, we develop a novel penalized inverse-variance weighted (pIVW) estimator, which adjusts the original IVW estimator to account for the weak IV issue by using a penalization approach to prevent the denominator of the pIVW estimator from being close to zero. Moreover, we adjust the variance estimation of the pIVW estimator to account for the presence of balanced horizontal pleiotropy. We show that the recently proposed debiased IVW (dIVW) estimator is a special case of our proposed pIVW estimator. We further prove that the pIVW estimator has smaller bias and variance than the dIVW estimator under some regularity conditions. We also conduct extensive simulation studies to demonstrate the performance of the proposed pIVW estimator. Furthermore, we apply the pIVW estimator to estimate the causal effects of five obesity-related exposures on three coronavirus disease 2019 (COVID-19) outcomes. Notably, we find that hypertensive disease is associated with an increased risk of hospitalized COVID-19; and peripheral vascular disease and higher body mass index are associated with increased risks of COVID-19 infection, hospitalized COVID-19 and critically ill COVID-19.

## 1. Introduction

It is of scientific interest to estimate the causal effects of modifiable risk factors on various health outcomes in epidemiological studies. For example, estimating the causal effects of modifiable risk factors on the coronavirus disease 2019 (COVID-19) outcomes is currently one of the most pressing global public health problems (Jordan et al., 2020; Zheng et al., 2020). The COVID-19 pandemic, caused by severe acute respiratory syndrome coronavirus 2 (SARS-CoV-2), has posed a serious threat to human health all over the world (Pascarella et al., 2020). It is crucial to identify causal risk factors associated with COVID-19 incidence and mortality so that we can develop more effective prevention and intervention strategies. One major challenge is the unmeasured confounding bias for the exposure-outcome relationship in observational epidemiological studies.

To address this challenge, Mendelian randomization (MR) utilizes genetic variants as instrumental variables (IVs) to estimate the causal effect of an exposure variable on an outcome of interest even in the presence of unmeasured confounders (Smith and Ebrahim, 2003, 2004; Sheehan et al., 2008). With the increasing availability of summary-level data from genome-wide association studies (GWASs), many MR methods have been developed based on GWAS summary-level data (Lawlor, 2016; Zheng et al., 2017). However, the validity of MR analysis critically depends on the following three core assumptions defining a valid IV (Didelez and Sheehan, 2007; Lawlor et al., 2008):

1. IV relevance: the IV must be associated with the exposure;
2. IV independence: the IV is independent of any confounder of the exposure-outcome relationship;
3. Exclusion restriction: the IV affects the outcome only through the exposure.

When any one of these three IV assumptions is violated, conventional MR analysis may yield biased estimation of the causal effect. In particular, the IV relevance assumption can be nearly violated when the IVs are only weakly associated with the exposure variable (Burgess and Thompson, 2011; Burgess et al., 2011; Davies et al., 2015). In MR studies, the weak IV bias may occur when the genetic variants only explain a small proportion of variance for the exposure variable. On the other hand, the widespread horizontal pleiotropy in human genome can also lead to the violation of the exclusion restriction assumption (Verbanck et al., 2018; Hemani et al., 2018), which is a phenomenon that the genetic variants directly affect the outcome not mediated by the exposure variable (see Figure 1 for a graphical illustration).

**Figure 1.**
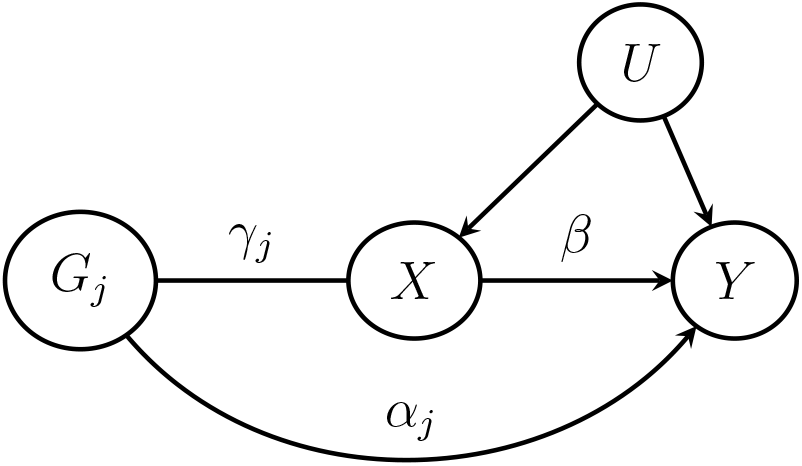
The relationships among the *j*th genetic variant *G*_*j*_, the exposure *X*, the outcome *Y* and the unmeasured confounder *U*. The effect of *G*_*j*_ on *X* is *γ*_*j*_, the direct effect (pleiotropic effect) of *G*_*j*_ on *Y* is *α*_*j*_, and the causal effect of *X* on *Y* is *β*.

The inverse-variance weighted (IVW) estimator is one of the most popular MR methods that has been widely used in health studies (Burgess et al., 2013). It has a simple and explicit expression, which combines the estimated causal effects from multiple IVs into a weighted average with the idea borrowed from the fixed-effect meta-analysis literature (Brockwell and Gordon, 2001). Despite its widespread popularity, recent studies pointed out that the IVW estimator can be seriously biased in the presence of weak IVs (Zhao et al., 2020; Ye et al., 2021). MR-RAPS is a maximum profile likelihood estimator, which was shown to be robust to weak IVs (Zhao et al., 2020). However, MR-RAPS has no closed-form solution and might not have unique estimates. Recently, the debiased IVW (dIVW) estimator was proposed to account for the weak IV issue by a simple modification to the IVW estimator (Ye et al., 2021). The dIVW estimator has been proved to be consistent even in the presence of many weak IVs under certain conditions. Nevertheless, as a ratio estimator, the dIVW estimator is still likely to yield a biased estimate when its denominator is close to zero. In fact, when the denominator is close to zero, a ratio estimator may have a heavy-tailed distribution and thus may not even have finite moments (Press, 1969; Piegorsch and Casella, 1985).

In this article, we develop a novel penalized inverse-variance weighted (pIVW) estimator, where the original IVW estimator is adjusted by a proposed penalized log-likelihood function. Through the penalization, we can prevent the denominator in the ratio estimator from being close to zero and thus provide improved estimation in the presence of weak IVs. Moreover, we account for the balanced horizontal pleiotropy by adjusting the variance estimation of the pIVW estimator. The proposed pIVW estimator has some attractive features. First, our theoretical and numerical results show that the proposed pIVW estimator has smaller bias and variance than the dIVW estimator under some regularity conditions. Second, it is consistent and asymptotically normal even in the presence of many weak IVs, and requires no more assumptions than the dIVW estimator. Third, it has a unique and closed-form solution, whereas some other robust MR methods (e.g., MR-RAPS) do not have a closed-form solution and might not have unique estimates in practice.

We demonstrate the improved performance of the proposed pIVW estimator compared to the other competing MR methods via extensive simulation studies. Furthermore, we apply the pIVW estimator to estimate the causal effects of five obesity-related exposures (i.e., peripheral vascular disease, dyslipidemia, hypertensive disease, type 2 diabetes and body mass index (BMI)) on three COVID-19 outcomes (i.e., COVID-19 infection, hospitalized COVID-19 and critically ill COVID-19). We find that hypertensive disease is significantly associated with an increased risk of hospitalized COVID-19; and peripheral vascular disease and higher BMI are significantly associated with increased risks of COVID-19 infection, hospitalized COVID-19 and critically ill COVID-19.

## 2. The Two-Sample MR Design and Prior Work

### 2.1 Linear Structural Models

Suppose that there are *p* independent genetic variants 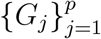. When there is no horizontal pleiotropy, the relationships among the genetic variants *G*_*j*_, the exposure *X*, the outcome *Y* and the unmeasured confounder *U* (as in Figure 1) can be formulated by the linear structural models as follows (Bowden et al., 2015):

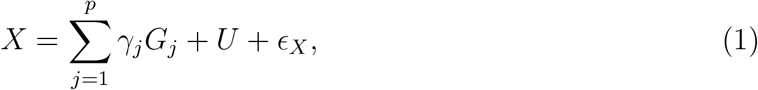

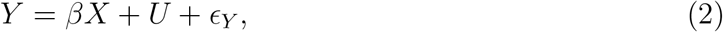

where *γ*_*j*_ is the genetic effect of *G*_*j*_ on *X, β* is the causal effect of our interest, and *ϵ*_*X*_ and *ϵ*_*Y*_ are mutually independent random errors. Let Γ_*j*_ denotes the effect of *G*_*j*_ on *Y*, then we have Γ_*j*_ = *βγ*_*j*_ by substituting Equation (1) for *X* in Equation (2).

Let 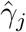 and 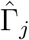 be the estimates of *γ*_*j*_ and Γ_*j*_ with the variances 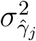 and 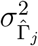, respectively. In the two-sample MR design, 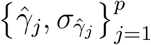 and 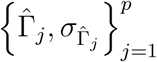 can be obtained from two independent GWASs (Lawlor, 2016). Since the GWASs generally involve large sample sizes, it is common to assume that 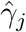 and 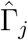 are independently distributed as 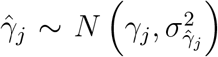 and 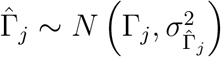, with known 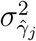 and 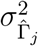, respectively (Zhao et al., 2020).

### 2.2 The IVW Estimator and Debiased IVW (dIVW) Estimator

The popular inverse-variance weighted estimator combines the estimated causal effects 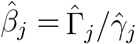 from multiple genetic variants with the weights 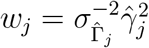 as follows (Burgess et al., 2013):

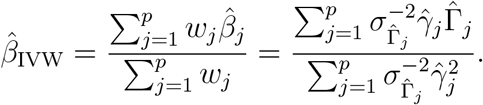

Let 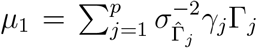 and 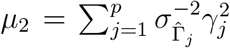. We have *β* = *μ*_1_*/μ*_2_ since Γ_*j*_ = *βγ*_*j*_ under models (1)-(2). As shown by Zhao et al. (2020), the IVW estimator can be approximated by

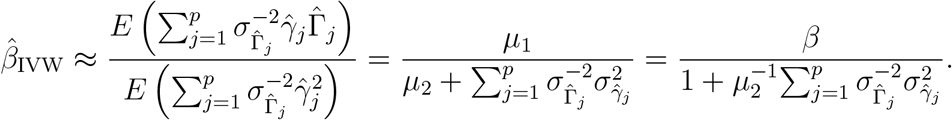

When there is no measurement error for *γ*_*j*_ (i.e., 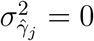), we have 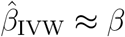. However, some recent studies have shown that the IVW estimator can be seriously biased toward zero for ignoring the measurement errors of *γ*_*j*_ especially in the presence of many weak IVs that have small 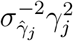 (Zhao et al., 2020; Ye et al., 2021).

To handle the bias due to weak IVs, the debiased IVW (dIVW) estimator (Ye et al., 2021) replaces the denominator in the IVW estimator by an unbiased estimator 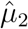 of *μ*_2_ as

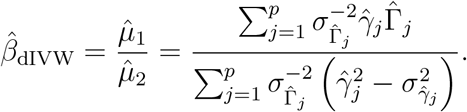

The dIVW estimator has been shown to be consistent and asymptotically normal under weaker conditions than the IVW estimator. However, we find that the dIVW estimator is more likely to yield extreme estimates in the presence of weak IVs (as shown in Web Figure 1 under the simulation study in Section 4). It can be shown that the denominator of the dIVW estimator has the same variance as the denominator of the IVW estimator, but the expectation of the former is closer to zero than that of the latter (see Web Appendix A for details). Because zero is a singular point for the denominator of a ratio, it may result in the extreme estimates of the dIVW estimator in the presence of weak IVs. To overcome the limitations of the IVW estimator and the dIVW estimator, we adjust the IVW estimator to account for the weak IVs by using a penalized log-likelihood function for *μ*_1_ and *μ*_2_, which can prevent the estimator of *μ*_2_ from being close to zero.

## 3. Method

### 3.1 The Penalized IVW (pIVW) Estimator

Assume that the estimators 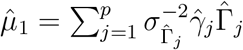 and 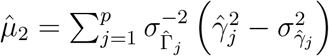 jointly follow the following bivariate normal distribution

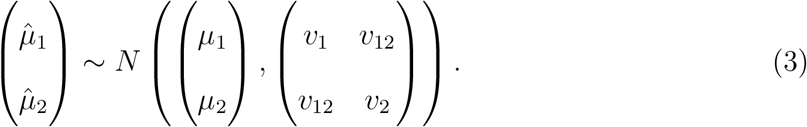

We propose the following penalized log-likelihood function to adjust the estimates of *μ*_1_ and *μ*_2_

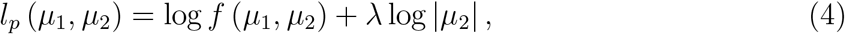

where *f* (*μ*_1_, *μ*_2_) denotes the bivariate normal density function of 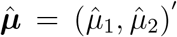 and *λ* > 0 is the penalty parameter. The penalized log-likelihood *l*_*p*_ (*μ*_1_, *μ*_2_) becomes small when *μ*_2_ approaches zero due to the penalty term *λ* log |*μ*_2_| (see Web Figure 2 for a graphical illustration). Therefore, an estimate of *μ*_2_ being close to zero is less preferable by *l*_*p*_ (*μ*_1_, *μ*_2_). Specifically, we obtain the following estimators of *μ* _1_ and *μ* _2_ by maximizing *l*_*p*_ (*μ*_1_, *μ* _2_) with closed-form expressions:

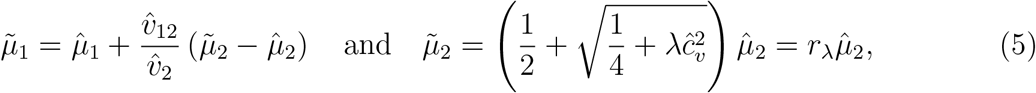

where 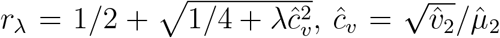 is the estimated coefficient of variation of 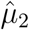, 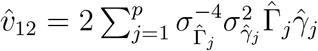 and 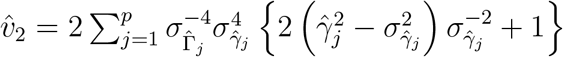 are the estimators of *v*_12_ and *v*_2_ respectively (see Web Appendix B for detailed derivation). Note that the bivariate normality assumption of 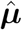 can be relaxed. In fact, we can take 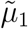 and 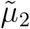 as the estimators that minimize the squared error loss function 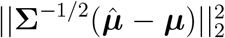 with a penalty term −2*λ* log |*μ*_2_|, that is,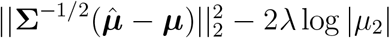, where ***μ*** and **Σ** denote the mean and the covariance matrix of 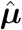 respectively. Then we propose the following penalized IVW (pIVW) estimator as a ratio of 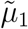 and 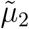

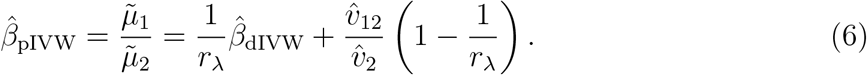

Note that, *r*_*λ*_ acts like a correction factor for 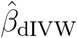. When the penalty parameter *λ* = 0, the correction factor *r*_*λ*_ = 1 and then 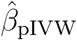 reduces to 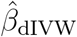. When *λ* > 0, we can see that *r*_*λ*_ > 1 and *r*_*λ*_ increases with the estimated coefficient of variation *ĉ*_*v*_. Therefore, when the estimated coefficient of variation *ĉ*_*v*_ is large (e.g., in the presence of many weak IVs), 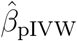 adjusts 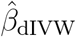 by *r*_*λ*_ to prevent the denominator 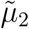 from being close to zero (see Web Figure 3 for the difference between 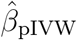 and 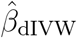 against various *r*_*λ*_). A good numerical example can be found in Section 5 (Table 4), where 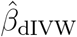 yields an extreme estimate of the causal effect of peripheral vascular disease on hospitalized COVID-19, and 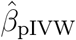 adjusts it by the correction factor *r*_*λ*_ in the case where no IV selection is performed.

To study the asymptotic properties of the pIVW estimator, we make the following Assumptions 1 and 2, which were also required for the consistency of the dIVW estimator (Ye et al., 2021).

#### Assumption 1

The number of IVs *p* diverges to infinity.

#### Assumption 2

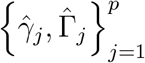 are independently distributed as 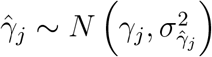 and 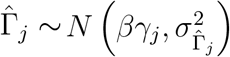 with known variances 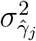 and 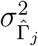. The ratio of variances 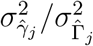 is bounded away from zero and infinity for all *j* = 1, …, *p*.

Assumptions 1 and 2 are reasonable in the two-sample MR design settings since the GWASs often have large sample sizes and a large number of genetic variants. The independence of 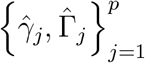 across genetic variants can be achieved by the linkage-disequilibrium clumping (Purcell et al., 2007).

Following Ye et al. (2021), we define the IV strength as

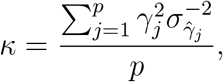

which is estimated by 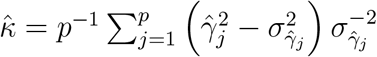. We also follow Ye et al. (2021) to define the effective sample size 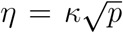. Note that the effective sample size *η* is determined by the IV strength and the number of IVs in the summary-level data, which is not the sample size of the original individual-level data in GWASs. Under Assumptions 1 and 2, it can be shown that 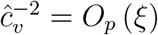 and 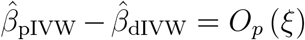, where 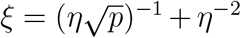 converges to zero as *η* → ∞. Therefore, given the consistency of 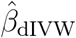, it is straightforward that 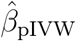 is also consistent as *η* → ∞. In the following Theorem 1 (a), we show that the bias of 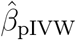 converges to zero at a faster rate than that of 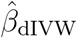 under the optimal *λ*_*opt*_ = 1. In Theorem 1 (b) together with Remark 2, we show that the variance of 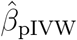 is smaller than that of 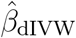 when *λ* > 0. In Theorem 1 (c), we also establish the asymptotic normality of 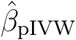, which requires no more assumptions comparing to 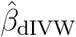.

#### Theorem 1

*Suppose that Assumptions 1-2 hold and the effective sample size η* → ∞. *Then, we have the following results:*

a. *The bias of* 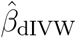 *is of order O* (*ξ*), *and the bias of* 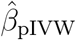 *is*

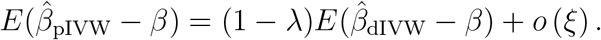 *The optimal λ*_*opt*_ = 1 *minimizes the absolute bias of* 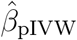, *which is only of order o* (*ξ*).
b. *The variances of* 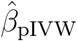 *and* 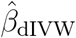 *are both of order O* (*ξ*). *But the difference between the variances of* 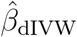 *and* 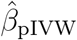 *is*

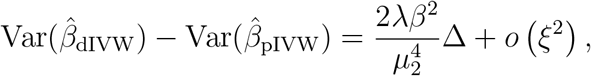

*where* 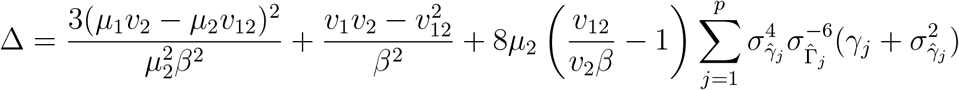.
c. *Further assume that* 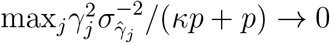. *Then*, 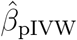 *is asymptotically normal*

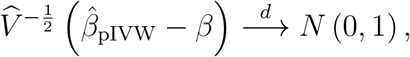

*where*

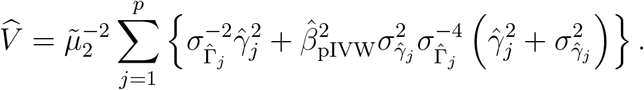

The proof of Theorem 1 is provided in Web Appendix C.

#### Remark 1

Theorem 1(a) states that 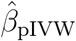 has smaller absolute bias than 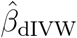 when 0 < *λ* < 2. In particular, the bias of 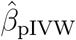 with *λ*_*opt*_ = 1 converges to zero at a faster rate than that of 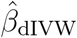.

#### Remark 2

Theorem 1(b) shows that 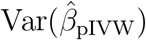 is smaller than 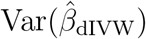 when Δ > 0. In fact, we have shown that Δ > 0 is generally true for complex traits, of which a single genetic variant can only explain a very small amount of total variances (Park et al., 2010; Shi et al., 2016; Boyle et al., 2017). More technical details can be found in Web Appendix D. Therefore, when *λ* > 0, 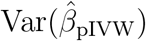 is smaller than 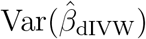 in MR settings. Together with Theorem 1(a), 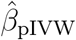 with *λ*_*opt*_ = 1 has smaller bias and variance than 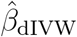.

#### Remark 3

In Theorem 1(c), we show that 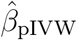 is asymptotically normal as *η* → ∞. Therefore, the confidence interval of *β* can be derived from the normal approximation of 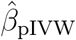. Alternatively, we can derive the confidence interval of *β* based on bootstrapping Fieller’s method (Fieller, 1954; Hwang and Hwang, 1995), which has been shown to have better coverage level than that based on the normal approximation. More details can be found in Web Appendix E.

In the setting of many weak IVs, we may have *κ* → 0 as *p* → ∞ because more weak IVs are likely to be included into the analysis as the number of IVs *p* increases, which may reduce the IV strength *κ*. The above theorem holds in this case as long as the effective sample size *η* → ∞, which means that it allows the presence of many weak IVs.

### 3.2 Selection of Candidate Instruments

In this section, we extend Theorem 1 to the setting where IV selection is conducted to remove some weak IVs from the analysis, which is a common practice in MR studies to handle the weak IV bias.

Suppose that there is a selection dataset 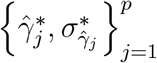 that is independent of the exposure and the outcome datasets. Then, an IV is included into the analysis when 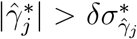 with a pre-set threshold *δ* > 0 (Zhao et al., 2019). Ye et al. (2021) showed that IV selection with an appropriate threshold *δ* could reduce the bias of the IVW estimator and improve the efficiency of the dIVW estimator. They also recommended a threshold 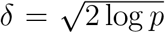 to guarantee a small probability of selecting any null IVs (i.e., *γ*_*j*_ = 0). When the IV selection is performed at a threshold *δ*, we follow Ye et al. (2021) to define the IV strength as

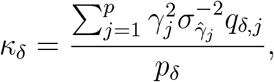

where 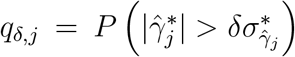 and 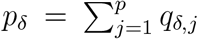. Let 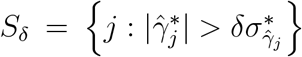 be the set of selected IVs, and 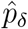 be the number of selected IVs within *S*_*δ*_. Then, we can estimate *κ*_*δ*_ by 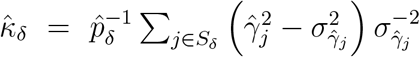. In the IV selection setting, we define the effective sample size 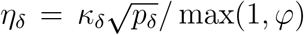 and 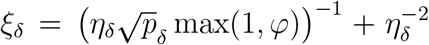, where 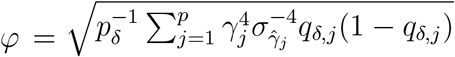. To study the theoretical properties of the proposed pIVW estimator under IV selection, we have the following Assumption 3 for the summary-level data in the selection dataset.

#### Assumption 3

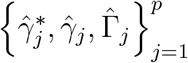 are mutually independent and 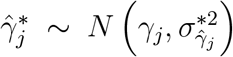 with known variance 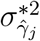 for every *j*. The ratio of variances 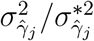 is bounded away from zero and infinity for all *j* = 1, …, *p*.

Given a selection threshold *δ*, we evaluate the dIVW estimator 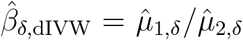 and the proposed pIVW estimator 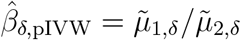 using the selected IVs within the set *S*_*δ*_. Under Assumptions 1-3, we have 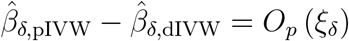. The following Theorem 2 shows that the asymptotic properties of the pIVW estimator in Theorem 1 still hold under the IV selection as the effective sample size *η*_*δ*_ → ∞.

#### Theorem 2

*Suppose that Assumptions 1-3 hold and the effective sample size η*_*δ*_ → ∞. *Then, we have the following results:*

a. *The bias of* 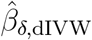 *is of order O* (*ξ*_*δ*_), *and the bias of* 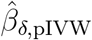 *is*

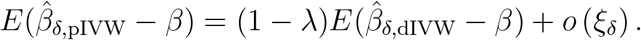 *The optimal λ*_*opt*_ = 1 *minimizes the absolute bias of* 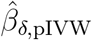, *which is only of order o* (*ξ*_*δ*_).
b. *The variances of* 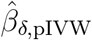 *and* 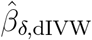 *are both of order O* (*ξ*_*δ*_). *But the difference between the variances of* 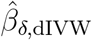 *and* 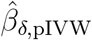 *is*

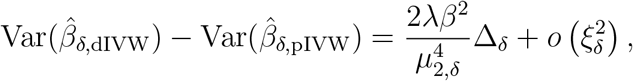

*where* 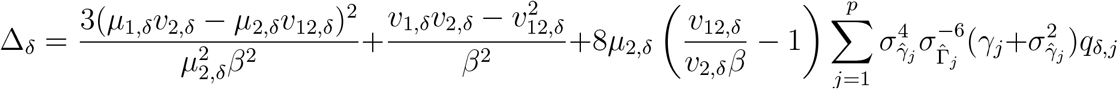.
c. *Further assume that* 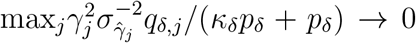. *Then*, 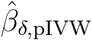 *is asymptotically normal*

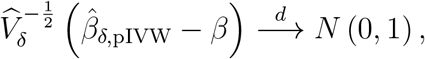

*where*

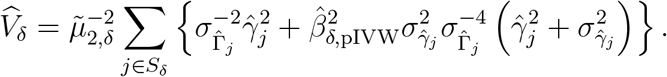

The proof of Theorem 2 is provided in Web Appendix F. Theorem 2 shows that 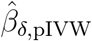 has smaller absolute bias than 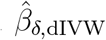 when 0 < *λ* < 2. The bias of 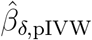 with the optimal *λ*_*opt*_ = 1 converges to zero faster than that of 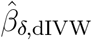. We also prove that Δ_*δ*_ > 0 generally holds in the genetic studies, and therefore 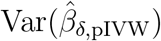 is smaller than 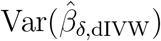 when *λ* > 0. The pIVW estimator is still consistent and asymptotically normal after accounting for the IV selection. We extend the results for the dIVW estimator in Ye et al. (2021) to the pIVW estimator. Note that, the independent datasets for IV selection might not be available for some traits in practice. However, the pIVW estimator is still useful in this case, since it can handle the weak IV bias even without IV selection as shown in Theorem 1.

### 3.3 Accounting for Balanced Horizontal Pleiotropy

When there exists horizontal pleiotropy (i.e., non-zero direct effect of *G*_*j*_ on *Y* not mediated by *X*), the linear structural model (2) can be modified as follows (Bowden et al., 2015):

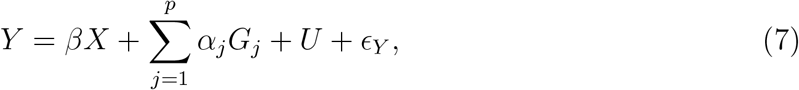

where *α*_*j*_ denotes the direct genetic effect of *G*_*j*_ on the outcome *Y* (i.e., pleiotropic effect). In this case, we have Γ_*j*_ = *βγ*_*j*_ + *α*_*j*_. We follow a common practice in many MR methods to assume that the horizontal pleiotropy is balanced (i.e., the pleiotropic effect has mean zero) and treat *α*_*j*_ as random effect following *α*_*j*_ ∼ *N* (0, *τ* ^2^) (Bowden et al., 2017; Zhao et al., 2020; Ye et al., 2021). Then, we have 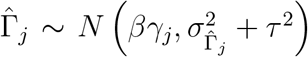 in the presence of balanced horizontal pleiotropy. To account for the balanced horizontal pleiotropy, we estimate the variance of 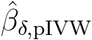 by

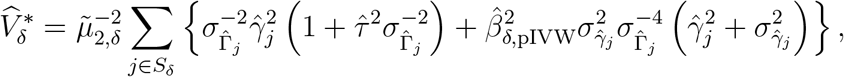

where we follow Ye et al. (2021) to derive the estimator of *τ* ^2^ as

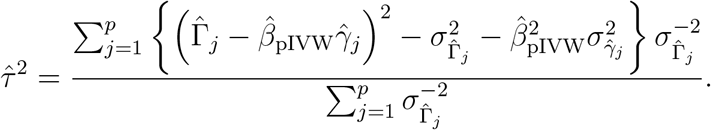

To establish the theoretical results for the pIVW estimator in the presence of balanced horizontal pleiotropy, we replace 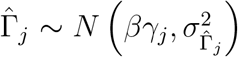 by 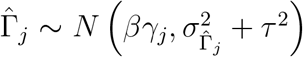 in Assumption 2, and assume that 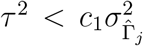 with a constant *c*_1_ > 0 for all *j*. We further assume that 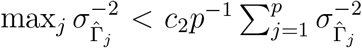 for a constant *c*_2_ > 0 in Theorems 1 (c) and 2 (c). Then, Theorems 1 and 2 can be extended to the situation with balanced horizontal pleiotropy. The proofs are provided in Web Appendices C and F respectively.

## 4. Simulation Study

### 4.1 Simulation Settings

We generate the summary-level data for 1000 IVs from 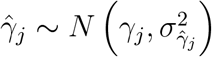 and 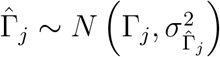 independently. For the true *γ*_*j*_, we consider a scenario with many weak IVs and many null IVs as in Ye et al. (2021), where we randomly generate *γ*_*j*_ ∼ *N* (0, 0.02^2^) for the weak IVs and let *γ*_*j*_ = 0 for the null IVs. We set the proportion of null IVs to be 95%, 90% and 80% corresponding to the effective sample size *η* around 4.33, 9.52 and 21.85, respectively. Then, we let Γ_*j*_ = *βγ*_*j*_ + *α*_*j*_, where *α*_*j*_ ∼ *N* (0, *τ* ^2^). We set *β* = 0.5, and *τ* = 0 and 0.01 which represent the absence and the presence of balanced horizontal pleiotropy, respectively.

The variances 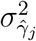 and 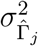 are given by 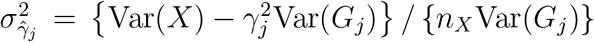 and 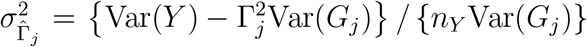, where *n*_*X*_ and *n*_*Y*_ denote the sample sizes of the GWASs for the exposure and the outcome, respectively. We set *n*_*X*_ = 0.5*n*_*Y*_ = 100, 000. For var(*G*_*j*_), we let *G*_*j*_ ∼ *Bin*(2, MAF_*j*_) and randomly generate the minor allele frequencies from MAF_*j*_ ∼ *U* (0.1, 0.5). For Var(*X*) and Var(*Y*), we calculate them from Equations (1) and (7) with the variances of *U, ϵ*_*X*_ and *ϵ*_*Y*_ being 2, respectively. Furthermore, we generate an independent dataset with 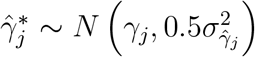 for the IV selection at threshold 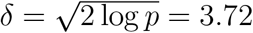. The simulation is based on 10,000 replicates.

We first investigate the impact of the penalty parameter *λ* on the performance of the proposed pIVW estimator, where *λ* increases from 0 to 2.5 by 0.5. Then, we compare the proposed pIVW estimator with *λ*_*opt*_ = 1 to other competing MR methods, including the IVW, the MR-Egger (Bowden et al., 2015), the MR-Median (Bowden et al., 2016), the MR-RAPS (Zhao et al., 2020) and the dIVW estimators. The performances among various methods are compared in terms of the relative bias (bias divided by the true *β*) and the empirical standard error of the estimated causal effect, as well as the coverage probability of nominal 95% confidence interval. For the pIVW etsimator, we present the coverage probability of bootstrapping Fieller’s confidence interval in this simulation. In Section 4.3, we also compare bootstrapping Fieller’s interval with the confidence interval derived from the normal approximation of the pIVW estimator under a wide range of parameter settings.

### 4.2 Simulation Results

The pIVW estimator has the smallest bias at *λ* = 1 as summarized in Table 1. The empirical standard error of the pIVW estimator decreases as *λ* increases. As the effective sample size *η* increases, the value of *λ* tends to have less influence on the performance of the pIVW estimator. We find similar results in the presence of balanced horizontal pleiotropy and IV selection (see Web Tables 1-3). Therefore, we recommend to choose the optimal *λ*_*opt*_ = 1 for the pIVW estimator in practice due to its smallest bias.

**Table 1.** The pIVW estimator with various penalty parameter λ. The true causal effect β = 0.5. No horizontal pleiotropy exists (τ = 0). No IV selection is conducted. The simulation is based on 10,000 replicates. Bias (%): bias divided by β; SE: empirical standard error; CP (%): coverage probability of the 95% confidence interval.

| $\eta$ | $\lambda$ | Bias | SE | CP |
| --- | --- | --- | --- | --- |
| 4.33 | 0 | 22.4 | 2.156 | 94.8 |
|  | 0.5 | 3.7 | 0.341 | 94.3 |
|  | 1 | -3.3 | 0.293 | 94.0 |
|  | 1.5 | -8.3 | 0.267 | 93.4 |
|  | 2 | -12.3 | 0.248 | 92.8 |
|  | 2.5 | -15.6 | 0.234 | 91.9 |
| 9.52 | 0 | 3.0 | 0.148 | 94.8 |
|  | 0.5 | 1.3 | 0.143 | 94.6 |
|  | 1 | -0.2 | 0.139 | 94.6 |
|  | 1.5 | -1.6 | 0.136 | 94.4 |
|  | 2 | -2.9 | 0.133 | 94.2 |
|  | 2.5 | -4.2 | 0.130 | 93.9 |
| 21.85 | 0 | 0.7 | 0.069 | 94.6 |
|  | 0.5 | 0.4 | 0.068 | 94.5 |
|  | 1 | 0.0 | 0.068 | 94.5 |
|  | 1.5 | -0.3 | 0.067 | 94.4 |
|  | 2 | -0.7 | 0.067 | 94.3 |
|  | 2.5 | -1.0 | 0.067 | 94.1 |

We next compare the pIVW estimator (*λ*_*opt*_ = 1) against the other five competing MR methods under the situations without horizontal pleiotropy and IV selection with the results summarized in Table 2. The pIVW estimator has negligible bias which is the smallest among all six methods. In contrast, the IVW, the MR-Egger and the MR-Median estimators have serious biases and poor coverage probabilities. The MR-RAPS and the dIVW estimators have relatively large empirical standard errors when the effective sample size *η* is small (*η* = 4.33), and we find that they yield some extreme estimates in this case (see Web Figure 1). As *η* increases, the differences in the performance among the MR-RAPS, the dIVW and the pIVW estimators become smaller.

**Table 2.** Comparison of the pIVW estimator (λ_opt_ = 1) with other competing MR methods. The true causal effect β = 0.5. No horizontal pleiotropy exists (τ = 0). No IV selection is conducted. The simulation is based on 10,000 replicates. Bias (%): bias divided by β; SE: empirical standard error; CP (%): coverage probability of the 95% confidence interval.

| $\eta$ | Method | Bias | SE | CP |
| --- | --- | --- | --- | --- |
| 4.33 | IVW | -88.0 | 0.028 | 0.0 |
|  | MR-Egger | -80.2 | 0.045 | 0.0 |
|  | MR-Median | -83.1 | 0.041 | 0.0 |
|  | MR-RAPS | 7.4 | 0.743 | 93.5 |
|  | dIVW | 22.4 | 2.156 | 94.6 |
|  | pIVW | -3.3 | 0.293 | 94.0 |
| 9.52 | IVW | -76.9 | 0.027 | 0.0 |
|  | MR-Egger | -63.5 | 0.042 | 0.0 |
|  | MR-Median | -67.6 | 0.040 | 0.0 |
|  | MR-RAPS | 1.2 | 0.120 | 94.7 |
|  | dIVW | 3.0 | 0.148 | 95.5 |
|  | pIVW | -0.2 | 0.139 | 94.6 |
| 21.85 | IVW | -59.1 | 0.024 | 0.0 |
|  | MR-Egger | -42.0 | 0.036 | 0.0 |
|  | MR-Median | -46.7 | 0.036 | 0.0 |
|  | MR-RAPS | 0.3 | 0.060 | 94.8 |
|  | dIVW | 0.7 | 0.069 | 94.8 |
|  | pIVW | 0.0 | 0.068 | 94.5 |

The results with IV selection at threshold 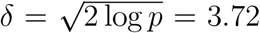 are given in Table 3. The proposed pIVW estimator still has the smallest bias among six methods and has smaller empirical standard error than the dIVW estimator. We obtain similar results in the presence of balanced horizontal pleiotropy (see Web Tables 4-5).

**Table 3.** Comparison of the pIVW estimator (λ_opt_ = 1) with other competing MR methods. The true causal effect β = 0.5. No horizontal pleiotropy exists (τ = 0). The IV selection threshold 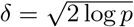. The simulation is based on 10,000 replicates. Bias (%): bias divided by β; SE: empirical standard error; CP (%): coverage probability of the 95% confidence interval.

| $\eta_\delta$ | Method | Bias | SE | CP |
| --- | --- | --- | --- | --- |
| 6.76 | IVW | -7.3 | 0.122 | 93.4 |
|  | MR-Egger | -41.9 | 0.376 | 89.3 |
|  | MR-Median | -11.6 | 0.140 | 95.1 |
|  | MR-RAPS | 1.4 | 0.137 | 95.8 |
|  | dIVW | 3.2 | 0.144 | 96.0 |
|  | pIVW | -0.1 | 0.136 | 95.3 |
| 10.26 | IVW | -7.9 | 0.079 | 91.4 |
|  | MR-Egger | -43.6 | 0.207 | 79.2 |
|  | MR-Median | -12.2 | 0.097 | 93.0 |
|  | MR-RAPS | 0.7 | 0.088 | 95.5 |
|  | dIVW | 1.4 | 0.090 | 95.5 |
|  | pIVW | 0.1 | 0.088 | 95.1 |
| 17.84 | IVW | -7.9 | 0.049 | 87.4 |
|  | MR-Egger | -48.6 | 0.130 | 51.2 |
|  | MR-Median | -12.5 | 0.062 | 88.3 |
|  | MR-RAPS | 0.4 | 0.055 | 94.9 |
|  | dIVW | 0.7 | 0.056 | 95.0 |
|  | pIVW | 0.2 | 0.055 | 94.7 |

We conduct an additional simulation study to mimic the individual-level data-generating mechanisms in GWASs. We first simulate the individual-level data based on the linear structural models (1) and (7). Then, we obtain the summary-level data by estimating the marginal effects and their standard errors from the linear regressions as in Ye et al. (2021) and Wang et al. (2022). We have similar findings in the simulation with individual-level data. More details are given in Web Appendix G and Web Tables 6-9.

### 4.3 Empirical Guidelines on η for Asymptotics

In Theorem 1, the asymptotic properties of 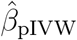 require the effective sample size *η* → ∞. To investigate how large of *η* is enough for the asymptotics, we conduct further simulations for 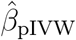 with *λ*_*opt*_ = 1 under a wide range of parameter settings, including: (1) *γ*_*j*_ ∼ *N* (0, *σ*^2^) with *σ* varying from 0.01 to 0.05, (2) the proportion of null IVs from 0 to 99%, and (3) *n*_*x*_ = *cn*_*y*_ = 100, 000 with *c* ranging from 0.1 to 10. The results show that the relative bias of 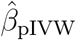 decreases more rapidly than that of 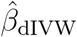 as *η* increases (see Figure 2 (a)). 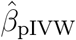 is nearly unbiased when *η* > 5, while 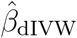 requires *η* > 15 to have a negligible bias. The variance of 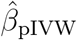 is much smaller than that of 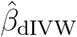 when *η* < 5, and they get close to each other as *η* increases (see Figure 2 (b)). The confidence interval derived from the normal approximation of 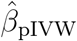 maintains nominal coverage probability when *η* > 10, while bootstrapping Fieller’s interval maintains nominal confidence probability when *η* > 5 (see Figure 2 (c)).

**Figure 2.**
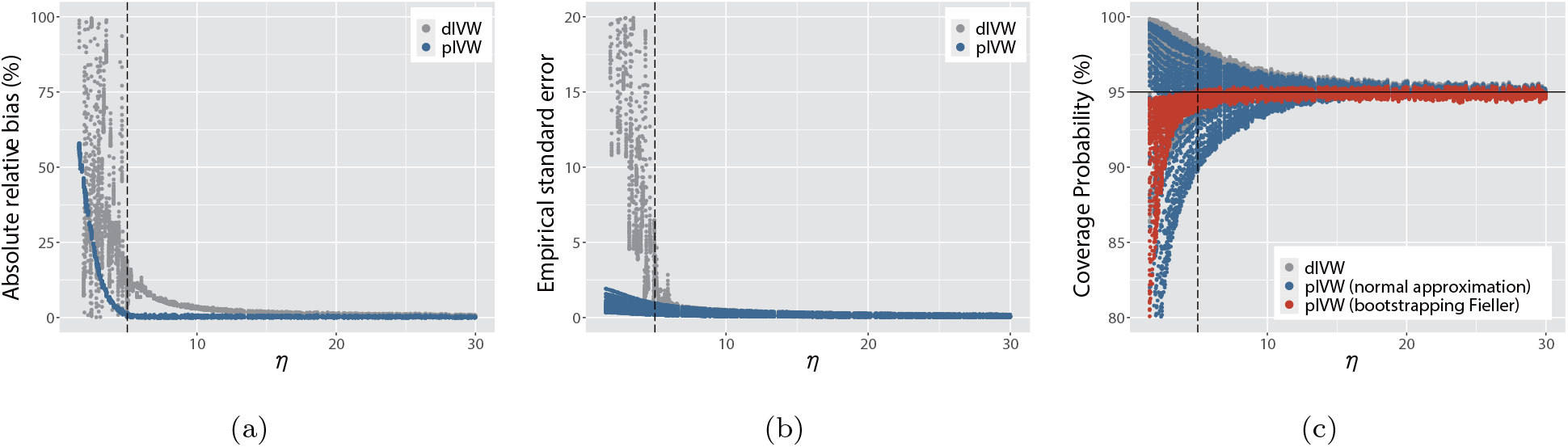
The plots of (a) the absolute relative biases (biases divided by *β*); (b) the empirical standard errors; and (c) the coverage probabilities of the 95% confidence intervals for the dIVW estimator and the pIVW estimator (*λ*_*opt*_ = 1) against the effective sample size *η*. The dashed line shows *η* = 5. The dots represent the simulation results under different settings of parameters based on 10,000 replicates. There is no horizontal pleiotropy (*τ* = 0) or IV selection. This figure appears in color in the electronic version of this article.

We have similar findings for *η*_*δ*_ under the IV selection (see Web Figure 4), where we consider a grid of two additional parameters, including the selection threshold *δ* from 1 to 4 and 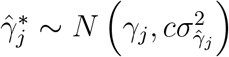 with *c* from 0.1 to 10. We also find similar results in the presence of balanced horizontal pleiotropy (see Web Figures 5-6). Therefore, we recommend that *η* (or *η*_*δ*_) should be larger than 5 for the pIVW estimator to have a negligible bias and a nominal coverage probability of the confidence interval. On the other hand, although the bias and variance of the pIVW estimator might not be negligible when *η* (or *η*_*δ*_) is less than 5, they are still much smaller than those of the dIVW estimator (see Figure 2 (a) and (b)).

## 5. Real Data Applications to COVID-19 Outcomes

In this section, we focus on estimating the causal effects of five obesity-related exposures (i.e., peripheral vascular disease, dyslipidemia, hypertensive disease, type 2 diabetes and BMI) on three COVID-19 outcomes: (1) COVID-19 infection, (2) hospitalized COVID-19 and (3) critically ill COVID-19 (COVID-19 Host Genetics Initiative, 2021). The GWAS summary-level data for the three COVID-19 outcomes is obtained from the COVID-19 Host Genetics Initiative (COVID-19 Host Genetics Initiative, 2020), which includes up to 49,562 cases and two million controls from 47 distinct studies. For BMI, the selection dataset is from Akiyama et al. (2017) with 173,430 individuals and the exposure dataset is from UK BioBank with 359,983 individuals (Abbott et al., 2018). For the other four obesity-related exposures, the selection datasets are from the GWAS meta-analysis of Genetic Epidemiology Research on Adult Health and Aging (GERA) with 53,991 individuals (Zhu et al., 2018), and the exposure datasets are from the GWAS meta-analysis of UK BioBank with 108,039 individuals (Zhu et al., 2018). More detailed data description is provided in Web Table 10. To exclude correlated IVs, we perform the linkage-disequilibrium clumping to remove the correlated genetic variants within 10Mb pairs and with the linkage disequilibrium *r*^2^ < 0.001. The numbers of IVs included into the analysis are from 1768 to 2338 for different datasets (see Web Table 11 for details).

From the results of the pIVW estimator with *λ*_*opt*_ = 1, we find significant positive causal effects of hypertensive disease on hospitalized COVID-19, and BMI on the three COVID-19 outcomes at significance level 0.05 (see Table 4 and Web Figures 7-8). Our findings agree with some recent epidemiological studies (Popkin et al., 2020; Nakeshbandi et al., 2020). Some previous MR studies have also found significant causal effects of BMI on the COVID-19 outcomes (Ponsford et al., 2020; Leong et al., 2021), but there is still no MR analysis on the hypertensive disease to the best of our knowledge. Additionally, the pIVW estimator suggests that peripheral vascular disease is significantly associated with higher risks of three COVID-19 outcomes under the IV selection at threshold 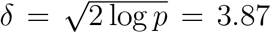. To our knowledge, there is a lack of MR studies about the associations between peripheral vascular disease and the COVID-19 outcomes, despite a high incidence of peripheral vascular disease in COVID-19 patients (Hanff et al., 2020). For type 2 diabetes and dyslipidemia, the pIVW estimator does not find any evidence of associations with the three COVID-19 outcomes. More results can be found in Web Table 11 and Web Figures 7-8.

**Table 4.** Estimated causal effects 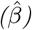 and estimated standard errors (SEs) of three obesity-related exposures (i.e., peripheral vascular disease (PVD), hypertensive disease (HD) and BMI) on the risk of hospitalized COVID-19. The pIVW estimator with the optimal λ_opt_ = 1

| Exposure | Method | No IV selection | | IV selection $\delta = \sqrt{2 \log p}$ | |
| --- | --- | --- | --- | --- | --- |
| | | $\hat{\eta}$ | $\hat{\beta}$ (SE) | $\hat{\eta}_{\delta}$ | $\hat{\beta}$ (SE) |
| PVD | IVW | 1.68 | 0.007 (0.013) | 3.98 | 0.233 (0.055) |
|  | MR-Egger |  | 0.022 (0.019) |  | 0.292 (0.071) |
|  | MR-Median |  | 0.032 (0.020) |  | 0.367 (0.098) |
|  | MR-RAPS |  | 0.379 (0.550) |  | 0.419 (0.217) |
|  | dIVW |  | 4.413 (95.942) |  | 0.670 (0.317) |
|  | pIVW |  | 0.219 (0.429) |  | 0.594 (0.245) |
| HD | IVW | 28.85 | 0.089 (0.033) | 12.04 | 0.135 (0.065) |
|  | MR-Egger |  | 0.068 (0.047) |  | 0.199 (0.091) |
|  | MR-Median |  | 0.109 (0.059) |  | 0.132 (0.099) |
|  | MR-RAPS |  | 0.241 (0.095) |  | 0.161 (0.080) |
|  | dIVW |  | 0.246 (0.093) |  | 0.163 (0.079) |
|  | pIVW |  | 0.244 (0.093) |  | 0.163 (0.078) |
| BMI | IVW | 218.93 | 0.382 (0.077) | 37.36 | 0.371 (0.100) |
|  | MR-Egger |  | 0.455 (0.105) |  | 0.563 (0.140) |
|  | MR-Median |  | 0.549 (0.141) |  | 0.361 (0.176) |
|  | MR-RAPS |  | 0.466 (0.097) |  | 0.397 (0.106) |
|  | dIVW |  | 0.468 (0.096) |  | 0.397 (0.106) |
|  | pIVW |  | 0.468 (0.096) |  | 0.397 (0.106) |

The other competing MR methods provide very different causal effect estimates when the estimated effective sample size 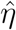 or 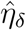 is small (see Table 4; see Web Appendix H for the estimation of 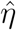 and 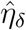). For peripheral vascular disease with a very small 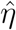 when no IV selection is performed, the IVW estimator has a very small estimate (0.007) which might be biased toward zero, because its denominator is a biased estimator of *μ*_2_ and might overestimate *μ*_2_ in the presence of many weak IVs (Ye et al., 2021). In contrast, the dIVW estimator yields a relatively large estimate (4.413) with an extreme estimated standard error (95.942), which possibly overestimates the causal effect due to the presence of many weak IVs. In this case, the pIVW estimator adjusts the dIVW estimator by the correction factor *r*_*λ*_ = 20.86, and provides an estimate being 0.219 with the estimated standard error being 0.429. After we perform IV selection to remove the weak IVs, the estimates from all the methods are in similar magnitudes. For BMI with large 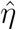 and 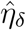, there is a smaller discrepancy among these methods, and the pIVW estimator and the dIVW estimator provide similar results in this case.

## 6. Discussion

The popular IVW estimator suffers from substantial bias in the presence of weak IVs, a common challenge in MR studies. In this paper, we develop a novel penalized IVW (pIVW) estimator to prevent the denominator of the ratio from being close to zero to reduce the bias due to the presence of many weak IVs. Moreover, we allow for the balanced horizontal pleiotropy by adjusting the variance estimation of the proposed pIVW estimator. Both simulation studies and real data analysis demonstrate the improved performance of the proposed pIVW estimator compared to the original IVW estimator and the recent dIVW estimator (Ye et al., 2021).

Our pIVW estimator has multiple advantages. First, our theoretical and numerical results show that the bias of the pIVW estimator with the optimal *λ*_*opt*_ = 1 converges to zero at a faster rate than that of the dIVW estimator as the effective sample size *η* (or *η*_*δ*_) increases. Meanwhile, the pIVW estimator with the optimal *λ*_*opt*_ = 1 has smaller variance than the dIVW estimator. Second, the proposed pIVW estimator is consistent and asymptotically normal even in the presence of many weak IVs, and requires no more assumptions than the dIVW estimator. The dIVW estimator can also be viewed as a special case of our proposed pIVW estimator, because the dIVW estimator is equivalent to the pIVW estimator with *λ* = 0. When *λ* > 0, their difference converges to zero as the effective sample size *η* (or *η*_*δ*_) increases. Third, the pIVW has a unique and closed-form solution, whereas many competing MR methods that are robust to the weak IVs do not have a closed-form solution and might have multiple numerical solutions (Zhao et al., 2019, 2020). In future work, we plan to extend the proposed penalization approach to other MR estimators to handle the weak IV bias, for instance, a penalized MR-Egger estimator (Bowden et al., 2015) to account for the unbalanced horizontal pleiotropy, and to account for the linkage disequilibrium (Wang et al., 2022).

## Data Availability

All data referred to in the manuscript is publicly available.

## Data Availability Statement

The GWAS summary data used in this article are available at the URLs as follows: the COVID-19 Host Genetics Initiative (release 5) https://www.covid19hg.org/results/r5/; the BMI selection data ftp://ftp.ebi.ac.uk/pub/databases/gwas/summary_statistics/AkiyamaM_28892062_GCST004904; the BMI exposure data http://www.nealelab.is/uk-biobank/; and four obesity-related diseases: the selection data (GERA data) and the exposure data (UK BioBank data) https://cnsgenomics.com/content/data.

## Supporting Information

The R package for the pIVW method including source code and example data is publicly available at https://github.com/siqixu/mr.pivw.

