## Supporting Information for "A Novel Penalized Inverse-Variance Weighted Estimator for Mendelian Randomization with Applications to COVID-19 Outcomes"

### Web Appendix A: Comparison of the Denominators for the IVW Estimator and the dIVW Estimator

Assume that the variances $\sigma_{\hat{\Gamma}_j}^2$ and $\sigma_{\hat{\gamma}_j}^2$ of $\hat{\Gamma}_j$ and $\hat{\gamma}_j$ are known, respectively, and $\hat{\gamma}_j$s are independently distributed as $\hat{\gamma}_j \sim N\left(\gamma_j, \sigma_{\hat{\gamma}_j}^2\right)$ for $j = 1, \dots, p$. Then, the expectation and variance of the denominator in the IVW estimator are respectively

$$E\left(\sum_{j=1}^{p} \sigma_{\hat{\Gamma}_j}^{-2} \hat{\gamma}_j^2\right) = \mu_2 + \sigma_{\hat{\Gamma}_j}^{-2} \sigma_{\hat{\gamma}_j}^2,$$

$$\text{Var}\left(\sum_{j=1}^{p} \sigma_{\hat{\Gamma}_j}^{-2} \hat{\gamma}_j^2\right) = \sum_{j=1}^{p} \sigma_{\hat{\Gamma}_j}^{-4} \left(4\sigma_{\hat{\gamma}_j}^2 \gamma_j^2 + 2\sigma_{\hat{\gamma}_j}^4\right).$$

The expectation and variance of the denominator in the dIVW estimator are respectively

$$E\left(\sum_{j=1}^{p} \sigma_{\hat{\Gamma}_j}^{-2} \left(\hat{\gamma}_j^2 - \sigma_{\hat{\gamma}_j}^2\right)\right) = \mu_2,$$

$$\text{Var}\left(\sum_{j=1}^{p} \sigma_{\hat{\Gamma}_j}^{-2} \left(\hat{\gamma}_j^2 - \sigma_{\hat{\gamma}_j}^2\right)\right) = \sum_{j=1}^{p} \sigma_{\hat{\Gamma}_j}^{-4} \left(4\sigma_{\hat{\gamma}_j}^2 \gamma_j^2 + 2\sigma_{\hat{\gamma}_j}^4\right).$$

### Web Appendix B: Derivation of the pIVW Estimator

The penalized log-likelihood function of $\mu_1$ and $\mu_2$ is

$$l_p(\mu_1, \mu_2) = -\frac{1}{2}\left(1 - \frac{v_{12}^2}{v_1 v_2}\right)^{-1} \left\{\frac{(\hat{\mu}_1 - \mu_1)^2}{v_1} - \frac{2v_{12}(\hat{\mu}_1 - \mu_1)(\hat{\mu}_2 - \mu_2)}{v_1 v_2} + \frac{(\hat{\mu}_2 - \mu_2)^2}{v_2}\right\} + \lambda \log|\mu_2| + C,$$

where $C$ is a constant unrelated to $\mu_1$ and $\mu_2$.

To maximize $l_p(\mu_1, \mu_2)$, we take its first derivative with respect to $\mu_1$ as follows,

$$\frac{\partial l_p(\mu_1, \mu_2)}{\partial \mu_1} = -\left(1 - \frac{v_{12}^2}{v_1 v_2}\right)^{-1} \left\{-\frac{\hat{\mu}_1 - \mu_1}{v_1} + \frac{v_{12}(\hat{\mu}_2 - \mu_2)}{v_1 v_2}\right\}.$$

From $\partial l_p(\mu_1, \mu_2)/\partial \mu_1$, we can see that $l_p(\mu_1, \mu_2)$ attains its maximum when

$$\mu_1 = \hat{\mu}_1 + \frac{v_{12}(\mu_2 - \hat{\mu}_2)}{v_2}.$$

By profiling out $\mu_1$, we get the following profile penalized log-likelihood function of $\mu_2$

$$l_p(\mu_2) = -\frac{(\hat{\mu}_2 - \mu_2)^2}{2v_2} + \lambda \log|\mu_2| + C.$$

The first derivative of $l_p(\mu_2)$ with respect to $\mu_2$ is

$$\frac{\partial l_p(\mu_2)}{\partial \mu_2} = \frac{\hat{\mu}_2 - \mu_2}{v_2} + \frac{\lambda}{\mu_2} = -\frac{(\mu_2 - \tilde{\mu}_{2,l})(\mu_2 - \tilde{\mu}_{2,r})}{\mu_2 v_2},$$

where

$$\tilde{\mu}_{2,l} = \frac{\hat{\mu}_2 - \sqrt{\hat{\mu}_2^2 + 4\lambda v_2}}{2} \text{ and } \tilde{\mu}_{2,r} = \frac{\hat{\mu}_2 + \sqrt{\hat{\mu}_2^2 + 4\lambda v_2}}{2}.$$

Since $\tilde{\mu}_{2,l} < 0 < \tilde{\mu}_{2,r}$, $l_p(\mu_2)$ increases when $\mu_2 \in (-\infty, \tilde{\mu}_{2,l}) \cup (0, \tilde{\mu}_{2,r})$ and decreases when $\mu_2 \in (\tilde{\mu}_{2,l}, 0) \cup (\tilde{\mu}_{2,r}, +\infty)$. Therefore, $l_p(\mu_2)$ can only attain its maximum at $\tilde{\mu}_{2,l}$ or $\tilde{\mu}_{2,r}$. Further, we have

$$l_p(\tilde{\mu}_{2,l}) = -\frac{\tilde{\mu}_{2,r}^2}{2v_2} + \lambda \log|\tilde{\mu}_{2,l}| + C,$$

$$l_p(\tilde{\mu}_{2,r}) = -\frac{\tilde{\mu}_{2,l}^2}{2v_2} + \lambda \log|\tilde{\mu}_{2,r}| + C.$$

When $\hat{\mu}_2 > 0$, we have $|\tilde{\mu}_{2,l}| < |\tilde{\mu}_{2,r}|$ and thus $l_p(\tilde{\mu}_{2,l}) < l_p(\tilde{\mu}_{2,r})$. When $\hat{\mu}_2 < 0$, we have $l_p(\tilde{\mu}_{2,l}) > l_p(\tilde{\mu}_{2,r})$. Hence, by maximizing $l_p(\mu_1, \mu_2)$, we obtain the following estimators of $\mu_1$ and $\mu_2$

$$\tilde{\mu}_1 = \hat{\mu}_1 + \frac{v_{12}(\tilde{\mu}_2 - \hat{\mu}_2)}{v_2},$$

$$\tilde{\mu}_2 = \frac{\hat{\mu}_2 + \mathrm{sign}(\hat{\mu}_2)\sqrt{\hat{\mu}_2^2 + 4\lambda v_2}}{2} = \left(\frac{1}{2} + \sqrt{\frac{1}{4} + \frac{\lambda v_2}{\hat{\mu}_2^2}}\right)\hat{\mu}_2.$$

Note that since the true values of $v_2$ and $v_{12}$ are unknown in practice, we replace them with their unbiased estimators $\hat{v}_2$ and $\hat{v}_{12}$ to obtain $\tilde{\mu}_1$ and $\tilde{\mu}_2$.

### Web Appendix C: Proof of Theorem 1

In the following proof, we write $a_n = O(b_n)$ if there exists a constant $c$ such that $|a_n| \leq cb_n$ for all $n$, $a_n = o(b_n)$ if $a_n/b_n \to 0$ as $n \to \infty$, and $a_n = \Theta(b_n)$ if there exists a constant $c$ such that $c^{-1}b_n \leq |a_n| \leq cb_n$ for all $n$. Let $\xrightarrow{p}$ denotes convergence in probability, and $\xrightarrow{d}$ denotes convergence in distribution. We write $X = O_p(Y)$ if $X/Y$ is bounded in probability, and $X = o_p(Y)$ if $X/Y \xrightarrow{p} 0$. The following proof of Theorem 1 takes into account the balanced horizontal pleiotropy ($\tau \neq 0$). The situation without horizontal pleiotropy can be considered as a special case with $\tau = 0$.

#### 1 Proof of Theorem 1 (a)

#### 1.1 Bias of dIVW Estimator

The dIVW estimator is

$$\hat{\beta}_{\mathrm{dIVW}} = \frac{\hat{\mu}_1}{\hat{\mu}_2} = \frac{\sum_{j=1}^{p} \sigma_{\hat{\Gamma}_j}^{-2} \hat{\gamma}_j \hat{\Gamma}_j}{\sum_{j=1}^{p} \sigma_{\hat{\Gamma}_j}^{-2} \left(\hat{\gamma}_j^2 - \sigma_{\hat{\gamma}_j}^2\right)}.$$

Let $\mu_1 = E(\hat{\mu}_1) = \beta \sum_{j=1}^{p} \sigma_{\hat{\Gamma}_j}^{-2} \gamma_j^2 = \Theta(\kappa p)$ and $\mu_2 = E(\hat{\mu}_2) = \sum_{j=1}^{p} \sigma_{\hat{\Gamma}_j}^{-2} \gamma_j^2 = \Theta(\kappa p)$ under Assumption 2. By Taylor series expansion, we have

$$\hat{\beta}_{\text{dIVW}} = \beta\ (\hat{x}_1 + 1)(\hat{x}_2 + 1)^{-1} = \beta + \beta\ (\hat{x}_1 - \hat{x}_2 + \hat{x}_2^2 - \hat{x}_1\hat{x}_2) + o_p(\xi),$$

where $\hat{x}_1 = \hat{\mu}_1/\mu_1 - 1$, $\hat{x}_2 = \hat{\mu}_2/\mu_2 - 1$, and $\xi = 1/\kappa p + 1/\kappa^2 p \to 0$ as the effective sample size $\eta = \kappa\sqrt{p} \to \infty$. Then, the bias of the dIVW estimator is

$$E\left(\hat{\beta}_{\text{dIVW}} - \beta\right) = \beta\left(\frac{v_2}{\mu_2^2} - \frac{v_{12}}{\mu_1\mu_2}\right) + o(\xi) = O(\xi),$$

where $v_2 = \text{Var}(\hat{\mu}_2) = \sum_{j=1}^{p} \sigma_{\hat{\Gamma}_j}^{-4}\left(4\sigma_{\hat{\gamma}_j}^2\gamma_j^2 + 2\sigma_{\hat{\gamma}_j}^4\right) = \Theta(\kappa p + p)$ and $v_{12} = \text{Cov}(\hat{\mu}_1, \hat{\mu}_2) = 2\beta \sum_{j=1}^{p} \sigma_{\hat{\Gamma}_j}^{-4} \sigma_{\hat{\gamma}_j}^2 \gamma_j^2 = \Theta(\kappa p)$ by Assumption 2.

**1.2 Bias of pIVW Estimator**

The pIVW estimator is

$$\hat{\beta}_{\text{pIVW}} = \frac{\hat{\mu}_1}{\tilde{\mu}_2} + \frac{\hat{v}_{12}}{\hat{v}_2}\left(1 - \frac{\hat{\mu}_2}{\tilde{\mu}_2}\right),$$

where $\tilde{\mu}_2 = \hat{\mu}_2\left(1 + \sqrt{1 + 4\lambda\hat{v}_2/\hat{\mu}_2^2}\right)/2$. The bias of the pIVW estimator is

$$E\left(\hat{\beta}_{\text{pIVW}} - \beta\right) = E\left(\frac{\hat{\mu}_1}{\tilde{\mu}_2} - \frac{\mu_1}{\mu_2}\right) + E\left(\frac{\hat{v}_{12}}{\hat{v}_2}\right)\left(1 - \frac{\hat{\mu}_2}{\tilde{\mu}_2}\right).$$

To evaluate $E\left(\frac{\hat{\mu}_1}{\tilde{\mu}_2} - \frac{\mu_1}{\mu_2}\right)$, we let $\tilde{x}_2 = \tilde{\mu}_2/m_2 - 1$ where $m_2 = E(\tilde{\mu}_2)$. By Taylor series expansion, we have

$$\frac{\hat{\mu}_1}{\tilde{\mu}_2} = \frac{\mu_1}{m_2}\ (\hat{x}_1 + 1)(\tilde{x}_2 + 1)^{-1} = \frac{\mu_1}{m_2} + \frac{\mu_1}{m_2}\ (\hat{x}_1 - \tilde{x}_2 + \tilde{x}_2^2 - \hat{x}_1\tilde{x}_2) + o_p(\xi).$$

Then,

$$E\left(\frac{\hat{\mu}_1}{\tilde{\mu}_2} - \frac{\mu_1}{\mu_2}\right) = \frac{\mu_1}{m_2} - \frac{\mu_1}{\mu_2} + \frac{\mu_1}{m_2}\{E(\tilde{x}_2^2) - E(\hat{x}_1\tilde{x}_2)\} + o(\xi). \quad \text{(A1)}$$

To obtain Equation (A1), we let $\mu_2^* = \mu_2\left(1 + \sqrt{1 + 4\lambda v_2/\mu_2^2}\right)/2$. By Taylor series expansion,

$$\frac{\tilde{\mu}_2}{\mu_2} = \frac{\mu_2^*}{\mu_2} + w\hat{x}_2 + \frac{\lambda v_2}{(2\mu_2^* - \mu_2)\mu_2}\hat{y}_2 + O_p(\xi^2) \quad \text{(A2)}$$

where $w = \mu_2^*/(2\mu_2^* - \mu_2) = 1 + O(\xi)$ and $\hat{y}_2 = \hat{v}_2/v_2 - 1$. Then, the expectation of $\tilde{\mu}_2/\mu_2$ is

$$\frac{m_2}{\mu_2} = \frac{\mu_2^*}{\mu_2} + o(\xi).$$

Since $\mu_2^* = \mu_2 + \lambda v_2/\mu_2^*$, we have

$$\frac{m_2 - \mu_2}{m_2} = \frac{\lambda v_2}{\mu_2^2} + o(\xi) = O(\xi).$$

Therefore,

$$\frac{\mu_1}{m_2} - \frac{\mu_1}{\mu_2} = \frac{\beta(\mu_2 - m_2)}{m_2} = -\frac{\beta\lambda v_2}{\mu_2^2} + o(\xi).$$

From Equation (A2), we have

$$E(\tilde{x}_2^2) = \frac{w^2 v_2}{m_2^2} + \frac{2\lambda \text{Cov}(\hat{\mu}_2, \hat{v}_2)}{\mu_2^3} + o(\xi^2) = \frac{v_2}{\mu_2^2} + o(\xi),$$

$$E(\hat{x}_1\tilde{x}_2) = \frac{w v_{12}}{\mu_1 m_2} + \frac{\lambda \text{Cov}(\hat{\mu}_1, \hat{v}_2)}{\beta\mu_2^3} + o(\xi^2) = \frac{v_{12}}{\mu_1\mu_2} + o(\xi),$$

where $\text{Cov}(\hat{\mu}_2, \hat{v}_2) = 8\sum_{j=1}^{p} \sigma_{\hat{\Gamma}_j}^{-6}\sigma_{\hat{\gamma}_j}^{4}\left(2\gamma_j^2 + \sigma_{\hat{\gamma}_j}^2\right) = \Theta(\kappa p + p)$ and $\text{Cov}(\hat{\mu}_1, \hat{v}_2) = 8\beta\sum_{j=1}^{p} \sigma_{\hat{\Gamma}_j}^{-6}\sigma_{\hat{\gamma}_j}^{4}\gamma_j^2 = \Theta(\kappa p)$. Then, Equation (A1) becomes

$$E\left(\frac{\hat{\mu}_1}{\tilde{\mu}_2} - \frac{\mu_1}{\mu_2}\right) = -\frac{\beta\lambda v_2}{\mu_2^2} + \beta\left(\frac{v_2}{\mu_2^2} - \frac{v_{12}}{\mu_1\mu_2}\right) + o(\xi). \quad \text{(A3)}$$

To evaluate $E\left(\frac{\hat{v}_{12}}{\hat{v}_2}\right)\left(1 - \frac{\hat{\mu}_2}{\tilde{\mu}_2}\right)$, we let $\hat{y}_{12} = \hat{v}_{12}/v_{12} - 1$. By Taylor series expansion,

$$\frac{\hat{v}_{12}}{\hat{v}_2} = \frac{v_{12}}{v_2}(\hat{y}_{12} + 1)(\hat{y}_2 + 1)^{-1} = \frac{v_{12}}{v_2}(1 + \hat{y}_{12} - \hat{y}_2 + \hat{y}_2^2 - \hat{y}_{12}\hat{y}_2) + o_p(\xi),$$

$$1 - \frac{\hat{\mu}_2}{\tilde{\mu}_2} = \frac{\lambda\hat{v}_2}{\tilde{\mu}_2^2} = \frac{\lambda v_2}{m_2^2}(\hat{y}_2 + 1)(\tilde{x}_2 + 1)^{-2} = \frac{\lambda v_2}{m_2^2}(1 + \hat{y}_2 - 2\tilde{x}_2) + O_p(\xi^2).$$

Then, we have

$$\frac{\hat{v}_{12}}{\hat{v}_2}\left(1 - \frac{\hat{\mu}_2}{\tilde{\mu}_2}\right) = \frac{\lambda v_{12}}{m_2^2}(\hat{y}_2 + 1 - 2\tilde{x}_2) + O_p(\xi^2),$$

and the expectation

$$E\left(\frac{\hat{v}_{12}}{\hat{v}_2}\right)\left(1 - \frac{\hat{\mu}_2}{\tilde{\mu}_2}\right) = \frac{\lambda v_{12}}{\mu_2^2} + o(\xi). \quad \text{(A4)}$$

Combining Equations (A3) and (A4), the bias of our proposed pIVW estimator is

$$\begin{aligned} E\left(\hat{\beta}_{\text{pIVW}} - \beta\right) &= E\left(\frac{\hat{\mu}_1}{\tilde{\mu}_2} - \frac{\mu_1}{\mu_2}\right) + E\left(\frac{\hat{v}_{12}}{\hat{v}_2}\right)\left(1 - \frac{\hat{\mu}_2}{\tilde{\mu}_2}\right) \\ &= -\frac{\beta\lambda v_2}{\mu_2^2} + \beta\left(\frac{v_2}{\mu_2^2} - \frac{v_{12}}{\mu_1\mu_2}\right) + \frac{\lambda v_{12}}{\mu_2^2} + o(\xi) \\ &= (1-\lambda)\beta\left(\frac{v_2}{\mu_2^2} - \frac{v_{12}}{\mu_1\mu_2}\right) + o(\xi). \end{aligned}$$

**2 Proof of Theorem 1 (b)**

By Taylor series expansion, we have

$$\begin{aligned} \left(\hat{\beta}_{\text{dIVW}} - \beta\right)^2 = \beta^2(&\hat{x}_1^2 + \hat{x}_2^2 - 2\hat{x}_1\hat{x}_2 - 2\hat{x}_1^2\hat{x}_2 + 4\hat{x}_1\hat{x}_2^2 - 2\hat{x}_2^3 + 3\hat{x}_1^2\hat{x}_2^2 - 6\hat{x}_1\hat{x}_2^3 + 3\hat{x}_2^4) \\ &+ o_p(\xi^2). \end{aligned}$$

Therefore,

$$E\left(\hat{\beta}_{\mathrm{dIVW}}-\beta\right)^2=\beta^2\left\{\frac{v_1}{\mu_1^2}+\frac{v_2}{\mu_2^2}-\frac{2v_{12}}{\mu_1\mu_2}-2E(\hat{x}_1^2\hat{x}_2)+4E(\hat{x}_1\hat{x}_2^2)-2E(\hat{x}_2^3)+3E(\hat{x}_1^2\hat{x}_2^2)\right.$$
$$\left.-6E(\hat{x}_1\hat{x}_2^3)+3E(\hat{x}_2^4)\right\}+o(\xi^2)=O(\xi), \qquad \text{(A5)}$$

where $v_1=\mathrm{Var}(\hat{\mu}_1)=\sum_{j=1}^{p}\sigma_{\hat{\Gamma}_j}^{-4}\left\{\left(\sigma_{\hat{\gamma}_j}^2\beta^2+\sigma_{\hat{\Gamma}_j}^2+\tau^2\right)\gamma_j^2+\left(\sigma_{\hat{\Gamma}_j}^2+\tau^2\right)\sigma_{\hat{\gamma}_j}^2\right\}=\Theta(\kappa p+p)$ by Assumption 2, and after some algebra, the expectations $E(\hat{x}_1^2\hat{x}_2)$, $E(\hat{x}_1\hat{x}_2^2)$, $E(\hat{x}_2^3)$, $E(\hat{x}_1^2\hat{x}_2^2)$, $E(\hat{x}_1\hat{x}_2^3)$ and $E(\hat{x}_2^4)$ are all of order $O(\xi^2)$. Since $E\left(\hat{\beta}_{\mathrm{dIVW}}-\beta\right)=O(\xi)$, the variance of the dIVW estimator is

$$\mathrm{Var}\left(\hat{\beta}_{\mathrm{dIVW}}\right)=E\left(\hat{\beta}_{\mathrm{dIVW}}-\beta\right)^2-E^2\left(\hat{\beta}_{\mathrm{dIVW}}-\beta\right)=O(\xi).$$

The difference of variance between $\hat{\beta}_{\mathrm{dIVW}}$ and $\hat{\beta}_{\mathrm{pIVW}}$ is

$$\mathrm{Var}\left(\hat{\beta}_{\mathrm{dIVW}}\right)-\mathrm{Var}\left(\hat{\beta}_{\mathrm{pIVW}}\right)$$
$$=E\left(\hat{\beta}_{\mathrm{dIVW}}-\beta\right)^2-E\left(\hat{\beta}_{\mathrm{pIVW}}-\frac{\mu_1}{m_2}\right)^2+E^2\left(\hat{\beta}_{\mathrm{pIVW}}-\frac{\mu_1}{m_2}\right)-E^2\left(\hat{\beta}_{\mathrm{dIVW}}-\beta\right).$$

First, we calculate $E^2\left(\hat{\beta}_{\mathrm{pIVW}}-\frac{\mu_1}{m_2}\right)-E^2\left(\hat{\beta}_{\mathrm{dIVW}}-\beta\right)$. Since

$$\beta-\frac{\mu_1}{m_2}=\beta\left(\frac{m_2-\mu_2}{m_2}\right)=\frac{\lambda\beta v_2}{\mu_2^2}+o(\xi),$$

we have

$$E^2\left(\hat{\beta}_{\mathrm{pIVW}}-\frac{\mu_1}{m_2}\right)=\left\{E\left(\hat{\beta}_{\mathrm{pIVW}}-\beta\right)+\left(\beta-\frac{\mu_1}{m_2}\right)\right\}^2$$
$$=\beta^2\left\{(1-\lambda)\left(\frac{v_2}{\mu_2^2}-\frac{v_{12}}{\mu_1\mu_2}\right)+\frac{\lambda v_2}{\mu_2^2}\right\}^2+o(\xi^2).$$

Therefore,

$$E^2\left(\hat{\beta}_{\mathrm{pIVW}}-\frac{\mu_1}{m_2}\right)-E^2\left(\hat{\beta}_{\mathrm{dIVW}}-\beta\right)=\frac{2\lambda\beta v_2 v_{12}}{\mu_2^4}+\frac{v_{12}^2}{\mu_2^4}(\lambda^2-2\lambda)+o(\xi^2). \qquad \text{(A6)}$$

Next, we calculate $E\left(\hat{\beta}_{\mathrm{dIVW}}-\beta\right)^2-E\left(\hat{\beta}_{\mathrm{pIVW}}-\frac{\mu_1}{m_2}\right)^2$. By Taylor series expansion, we have

$$\hat{\beta}_{\mathrm{pIVW}}-\frac{\mu_1}{m_2}=\frac{\hat{\mu}_1}{\tilde{\mu}_2}-\frac{\mu_1}{m_2}+\frac{\hat{v}_{12}}{\hat{v}_2}\left(1-\frac{\hat{\mu}_2}{\tilde{\mu}_2}\right)$$
$$=\frac{\mu_1}{m_2}(\hat{x}_1-\tilde{x}_2-\hat{x}_1\tilde{x}_2+\tilde{x}_2^2+\hat{x}_1\tilde{x}_2^2-\tilde{x}_2^3)+\frac{\lambda v_{12}}{m_2^2}(\hat{y}_2+1-2\tilde{x}_2)+O_p(\xi^2).$$

Then,

$$\left(\hat{\beta}_{\mathrm{pIVW}}-\frac{\mu_1}{m_2}\right)^2=\left(\frac{\mu_1}{m_2}\right)^2(\hat{x}_1^2+\tilde{x}_2^2-2\hat{x}_1\tilde{x}_2-2\hat{x}_1^2\tilde{x}_2+4\hat{x}_1\tilde{x}_2^2-2\tilde{x}_2^3+3\hat{x}_1^2\tilde{x}_2^2-6\hat{x}_1\tilde{x}_2^3$$
$$+3\tilde{x}_2^4)-\frac{2\lambda\mu_1 v_{12}}{m_2^3}(\hat{x}_1\hat{y}_2-\tilde{x}_2\hat{y}_2+\hat{x}_1-\tilde{x}_2-3\hat{x}_1\tilde{x}_2+3\tilde{x}_2^2)+\frac{\lambda^2 v_{12}^2}{m_2^4}$$
$$+o_p(\xi^2).$$

From Equation (A2), we have $E(\tilde{x}_1^2\tilde{x}_2) = E(\hat{x}_1^2\hat{x}_2) + o(\xi^2)$, $E(\tilde{x}_1\tilde{x}_2^2) = E(\hat{x}_1\hat{x}_2^2) + o(\xi^2)$, $E(\tilde{x}_2^3) = E(\hat{x}_2^3) + o(\xi^2)$, $E(\tilde{x}_1^2\tilde{x}_2^2) = E(\hat{x}_1^2\hat{x}_2^2) + o(\xi^2)$, $E(\tilde{x}_1\tilde{x}_2^3) = E(\hat{x}_1\hat{x}_2^3) + o(\xi^2)$, $E(\tilde{x}_2^4) = E(\hat{x}_2^4) + o(\xi^2)$, and $E(\tilde{x}_2\hat{y}_2) = \text{Cov}(\hat{\mu}_2, \hat{v}_2)/(\mu_2 v_2) + o(\xi)$. After some algebra, we have

$$
\begin{aligned}
E\left(\hat{\beta}_{\text{pIVW}} - \frac{\mu_1}{m_2}\right)^2 &\\
= &\left(\frac{\mu_1}{m_2}\right)^2\left(\frac{v_1}{\mu_1^2} + \frac{w^2 v_2}{m_2^2} - \frac{2wv_{12}}{\mu_1 m_2}\right) \\
&+ \beta^2\{-2E(\hat{x}_1^2\hat{x}_2) + 4E(\hat{x}_1\hat{x}_2^2) - 2E(\hat{x}_2^3) + 3E(\hat{x}_1^2\hat{x}_2^2) - 6E(\hat{x}_1\hat{x}_2^3) \\
&+ 3E(\hat{x}_2^4)\} + \frac{\lambda^2 v_{12}^2}{\mu_2^4} + \frac{6\lambda\beta v_{12}}{\mu_2^2}\left(\frac{v_2}{\mu_2^2} - \frac{v_{12}}{\mu_1\mu_2}\right) \\
&- 16\lambda\beta\left(\frac{v_{12}}{v_2} - \beta\right)\frac{\sum_{j=1}^{p}\sigma_{\hat{\Gamma}_j}^{-6}\sigma_{\hat{\gamma}_j}^{6}\left(\gamma_j^2\sigma_{\hat{\gamma}_j}^{-2} + 1\right)}{\mu_2^3} + o(\xi^2). \qquad \text{(A7)}
\end{aligned}
$$

From Equations (A5) and (A7), we have

$$
\begin{aligned}
E&\left(\hat{\beta}_{\text{dIVW}} - \beta\right)^2 - E\left(\hat{\beta}_{\text{pIVW}} - \frac{\mu_1}{m_2}\right)^2 \\
&= \beta^2\left(\frac{v_1}{\mu_1^2} + \frac{v_2}{\mu_2^2} - \frac{2v_{12}}{\mu_1\mu_2}\right) - \left(\frac{\mu_1}{m_2}\right)^2\left(\frac{v_1}{\mu_1^2} + \frac{w^2 v_2}{m_2^2} - \frac{2wv_{12}}{\mu_1 m_2}\right) - \frac{\lambda^2 v_{12}^2}{\mu_2^4} \\
&\quad - \frac{6\lambda\beta v_{12}}{\mu_2^2}\left(\frac{v_2}{\mu_2^2} - \frac{v_{12}}{\mu_1\mu_2}\right) + 16\lambda\beta\left(\frac{v_{12}}{v_2} - \beta\right)\frac{\sum_{j=1}^{p}\sigma_{\hat{\Gamma}_j}^{-6}\sigma_{\hat{\gamma}_j}^{6}\left(\gamma_j^2\sigma_{\hat{\gamma}_j}^{-2} + 1\right)}{\mu_2^3} + o(\xi^2) \\
&= \frac{2\lambda v_1 v_2}{\mu_2^4} + \frac{6\lambda\beta^2 v_2^2}{\mu_2^4} - \frac{14\lambda\beta v_2 v_{12}}{\mu_2^4} + \frac{(6\lambda - \lambda^2)v_{12}^2}{\mu_2^4} \\
&\quad + 16\lambda\beta\left(\frac{v_{12}}{v_2} - \beta\right)\frac{\sum_{j=1}^{p}\sigma_{\hat{\Gamma}_j}^{-6}\sigma_{\hat{\gamma}_j}^{6}\left(\gamma_j^2\sigma_{\hat{\gamma}_j}^{-2} + 1\right)}{\mu_2^3} + o(\xi^2), \qquad \text{(A8)}
\end{aligned}
$$

where we obtain the second equality by using $(m_2 - \mu_2)/m_2 = \lambda v_2/\mu_2^2 + o(\xi)$ and $1 - w = \lambda v_2/\mu_2^2 + o(\xi)$. Combining Equations (A6) and (A8), the difference in the variance between the dIVW estimator and the pIVW estimator is

$$
\begin{aligned}
\text{Var}\left(\hat{\beta}_{\text{dIVW}}\right) - \text{Var}\left(\hat{\beta}_{\text{pIVW}}\right) &= \frac{2\lambda v_1 v_2}{\mu_2^4} + \frac{6\lambda\beta^2 v_2^2}{\mu_2^4} - \frac{12\lambda\beta v_2 v_{12}}{\mu_2^4} + \frac{4\lambda v_{12}^2}{\mu_2^4} \\
&\quad + 16\lambda\beta\left(\frac{v_{12}}{v_2} - \beta\right)\frac{\sum_{j=1}^{p}\sigma_{\hat{\Gamma}_j}^{-6}\sigma_{\hat{\gamma}_j}^{6}\left(\gamma_j^2\sigma_{\hat{\gamma}_j}^{-2} + 1\right)}{\mu_2^3} + o(\xi^2) \\
&= \frac{2\lambda\beta^2}{\mu_2^4}\Delta + o(\xi^2),
\end{aligned}
$$

where

$$
\Delta = \frac{3(\mu_1 v_2 - \mu_2 v_{12})^2}{\mu_2^2\beta^2} + \frac{v_1 v_2 - v_{12}^2}{\beta^2} + 8\mu_2\left(\frac{v_{12}}{v_2\beta} - 1\right)\sum_{j=1}^{p}\sigma_{\hat{\Gamma}_j}^{-6}\sigma_{\hat{\gamma}_j}^{6}\left(\gamma_j^2\sigma_{\hat{\gamma}_j}^{-2} + 1\right).
$$

**3 Proof of Theorem 1 (c)**

We first prove

$$V^{-\frac{1}{2}}\left(\hat{\beta}_{\text{pIVW}}-\beta\right)=\left(\frac{\mu_2}{\sqrt{v}}\right)\frac{\hat{\mu}_1-\beta\hat{\mu}_2}{\tilde{\mu}_2}+\left(\frac{\mu_2}{\sqrt{v}}\right)\left(\frac{\hat{\mu}_2}{\tilde{\mu}_2}-1\right)\left(\beta-\frac{\hat{v}_{12}}{\hat{v}_2}\right)\xrightarrow{d}N(0,1),$$

where $V=\mu_2^{-2}\sum_{j=1}^{p}\left\{\sigma_{\hat{\Gamma}_j}^{-2}\left(\gamma_j^2+\sigma_{\hat{\gamma}_j}^2\right)\left(1+\tau^2\sigma_{\hat{\Gamma}_j}^{-2}\right)+\beta^2\sigma_{\hat{\gamma}_j}^2\sigma_{\hat{\Gamma}_j}^{-4}\left(\gamma_j^2+2\sigma_{\hat{\gamma}_j}^2\right)\right\}$ and $v=\mu_2^2V$. For this, we prove $\left(\frac{\mu_2}{\sqrt{v}}\right)\frac{\hat{\mu}_1-\beta\hat{\mu}_2}{\tilde{\mu}_2}\xrightarrow{d}N(0,1)$ and $\left(\frac{\mu_2}{\sqrt{v}}\right)\left(\frac{\hat{\mu}_2}{\tilde{\mu}_2}-1\right)\left(\beta-\frac{\hat{v}_{12}}{\hat{v}_2}\right)\xrightarrow{p}0$ in the following.

First, we prove $\tilde{\mu}_2/\mu_2\xrightarrow{p}1$. For this, we let $\tilde{\mu}_2=\hat{\mu}_2r_\lambda$ with $r_\lambda=\left(1+\sqrt{1+4\lambda\hat{v}_2/\hat{\mu}_2^2}\right)/2$. We have $\hat{\mu}_2/\mu_2\xrightarrow{p}1$ as $\eta\to\infty$, since $E(\hat{\mu}_2/\mu_2)=1$ and $\text{Var}(\hat{\mu}_2/\mu_2)=O(\xi)$. Similarly, $\hat{v}_2/v_2\xrightarrow{p}1$ as $\eta\to\infty$, since $E(\hat{v}_2/v_2)=1$ and $\text{Var}(\hat{v}_2/v_2)=O(\xi)$. Since $v_2/\mu_2^2=O(\xi)$, we have $r_\lambda\xrightarrow{p}1$ and thus $\tilde{\mu}_2/\mu_2\xrightarrow{p}1$ as $\eta\to\infty$.

Next, we prove $\frac{\hat{\mu}_1-\beta\hat{\mu}_2}{\sqrt{v}}\xrightarrow{d}N(0,1)$ as $p\to\infty$ by the Lindeberg Central Limit Theorem. Let

$$\frac{\hat{\mu}_1-\beta\hat{\mu}_2}{\sqrt{v}}=\frac{\sum_{j=1}^{p}\hat{x}_j}{\sqrt{\sum_{j=1}^{p}v_j}},$$

where $\hat{x}_j=\sigma_{\hat{\Gamma}_j}^{-2}\left\{\hat{\gamma}_j\hat{\Gamma}_j-\beta\left(\hat{\gamma}_j^2-\sigma_{\hat{\gamma}_j}^2\right)\right\}$ and $v_j=\sigma_{\hat{\Gamma}_j}^{-2}\left(\gamma_j^2+\sigma_{\hat{\gamma}_j}^2\right)\left(1+\tau^2\sigma_{\hat{\Gamma}_j}^{-2}\right)+\beta^2\sigma_{\hat{\gamma}_j}^2\sigma_{\hat{\Gamma}_j}^{-4}\left(\gamma_j^2+2\sigma_{\hat{\gamma}_j}^2\right)$. Let $\hat{z}_j=\hat{x}_j/\sqrt{v_j}$. Then, the Lindeberg's condition is

$$\frac{1}{v}\sum_{j=1}^{p}E\left(\hat{x}_j^2I\{|\hat{x}_j|>\epsilon\sqrt{v}\}\right)=\frac{1}{v}\sum_{j=1}^{p}E\left(v_j\hat{z}_j^2\,I\{\sqrt{v_j}|\hat{z}_j|>\epsilon\sqrt{v}\}\right)$$
$$\leq\max_j E\left(\hat{z}_j^2\,I\{\sqrt{v_j}|\hat{z}_j|>\epsilon\sqrt{v}\}\right).$$

By Assumption 2, we have $v=\Theta(\kappa p+p)$. Therefore, $\max_j v_j/v\to0$ as $\max_j\gamma_j^2\sigma_{\hat{\gamma}_j}^{-2}/(\kappa p+p)\to0$. Together with $E\left(\hat{z}_j^2\right)=1$, we verify the Lindeberg's condition $\frac{1}{v}\sum_{j=1}^{p}E\left(\hat{x}_j^2I\{|\hat{x}_j|>\epsilon\sqrt{v}\}\right)\to0$. Then, $\left(\frac{\mu_2}{\sqrt{v}}\right)\frac{\hat{\mu}_1-\beta\hat{\mu}_2}{\tilde{\mu}_2}\xrightarrow{d}N(0,1)$ follows from Slutsky's theorem.

Lastly, we prove $\left(\frac{\mu_2}{\sqrt{v}}\right)\left(\frac{\hat{\mu}_2}{\tilde{\mu}_2}-1\right)\left(\beta-\frac{\hat{v}_{12}}{\hat{v}_2}\right)\xrightarrow{p}0$. For this, we rewrite $\hat{\mu}_2/\tilde{\mu}_2-1$ as

$$\frac{\hat{\mu}_2}{\tilde{\mu}_2}-1=\frac{1-r_\lambda}{r_\lambda}=-\frac{\lambda\hat{v}_2}{\hat{\mu}_2^2}\frac{1}{r_\lambda^2},$$

where the last equality follows from $(r_\lambda-1)r_\lambda=\lambda\hat{v}_2/\hat{\mu}_2^2$. As such,

$$\left(\frac{\mu_2}{\sqrt{v}}\right)\left(\frac{\hat{\mu}_2}{\tilde{\mu}_2}-1\right)=-\left(\frac{\lambda v_2}{\sqrt{v}\mu_2}\right)\frac{\mu_2^2}{\hat{\mu}_2^2}\frac{\hat{v}_2}{v_2}\frac{1}{r_\lambda^2}=O_p\left(\sqrt{\xi}\right).$$

Together with $\beta-\frac{\hat{v}_{12}}{\hat{v}_2}=O_p(1)$, we have $\left(\frac{\mu_2}{\sqrt{v}}\right)\left(\frac{\hat{\mu}_2}{\tilde{\mu}_2}-1\right)\left(\beta-\frac{\hat{v}_{12}}{\hat{v}_2}\right)\xrightarrow{p}0$ as $\eta\to\infty$. Since $\left(\frac{\mu_2}{\sqrt{v}}\right)\frac{\hat{\mu}_1-\beta\hat{\mu}_2}{\tilde{\mu}_2}\xrightarrow{d}N(0,1)$ and $\left(\frac{\mu_2}{\sqrt{v}}\right)\left(\frac{\hat{\mu}_2}{\tilde{\mu}_2}-1\right)\left(\beta-\frac{\hat{v}_{12}}{\hat{v}_2}\right)\xrightarrow{p}0$, we get $V^{-\frac{1}{2}}\left(\hat{\beta}_{\text{pIVW}}-\beta\right)\xrightarrow{d}N(0,1)$ by Slutsky's theorem. Further, to prove that $\hat{V}^{-1/2}\left(\hat{\beta}_{\text{pIVW}}-\beta\right)\xrightarrow{d}N(0,1)$ holds with the estimator $\hat{V}$ of $V$, it suffices to prove $\tilde{\mu}_2/\mu_2 \xrightarrow{p} 1$ and $\hat{v}/v \xrightarrow{p} 1$. We have shown $\tilde{\mu}_2/\mu_2 \xrightarrow{p} 1$ above. Following Ye et al., 2021, it can be shown that $\hat{v}/v \xrightarrow{p} 1$ after replacing $\hat{\beta}_{\text{dIVW}}$ by $\hat{\beta}_{\text{pIVW}}$. Thus, we omit the details here.

### Web Appendix D: Proof of $\Delta > 0$

The following proof holds for both the situations with balanced horizontal pleiotropy ($\tau \neq 0$) and without horizontal pleiotropy ($\tau = 0$).

Let $\kappa_j = \gamma_j^2 \sigma_{\hat{\gamma}_j}^{-2}$. After some simple algebra, we have

$$\Delta = \frac{v_1 v_2}{\beta^2} + 3v_2^2 - \frac{6v_2 v_{12}}{\beta} + \frac{2v_{12}^2}{\beta^2} + 8\mu_2\left(\frac{v_{12}}{v_2\beta} - 1\right)\sum_{j=1}^{p} \sigma_{\hat{\Gamma}_j}^{-6}\sigma_{\hat{\gamma}_j}^{6}(\kappa_j + 1)$$

$$> v_2 \sum_{j=1}^{p} \sigma_{\hat{\Gamma}_j}^{-4}\sigma_{\hat{\gamma}_j}^{4}\kappa_j + 6v_2 \sum_{j=1}^{p} \sigma_{\hat{\Gamma}_j}^{-4}\sigma_{\hat{\gamma}_j}^{4} + 8\left(\sum_{j=1}^{p} \sigma_{\hat{\Gamma}_j}^{-4}\sigma_{\hat{\gamma}_j}^{4}\kappa_j\right)^2 - 8\sum_{j=1}^{p} \sigma_{\hat{\Gamma}_j}^{-2}\sigma_{\hat{\gamma}_j}^{2}\kappa_j \sum_{j=1}^{p} \sigma_{\hat{\Gamma}_j}^{-6}\sigma_{\hat{\gamma}_j}^{6}(\kappa_j + 1)$$

$$= 2\sum_{j=1}^{p} \sigma_{\hat{\Gamma}_j}^{-4}\sigma_{\hat{\gamma}_j}^{4}(2\kappa_j + 3)\sum_{j=1}^{p} \sigma_{\hat{\Gamma}_j}^{-4}\sigma_{\hat{\gamma}_j}^{4}(3\kappa_j + 2) - 8\sum_{j=1}^{p} \sigma_{\hat{\Gamma}_j}^{-2}\sigma_{\hat{\gamma}_j}^{2}\kappa_j \sum_{j=1}^{p} \sigma_{\hat{\Gamma}_j}^{-6}\sigma_{\hat{\gamma}_j}^{6}(\kappa_j + 1),$$

where we obtain the second inequality by using $v_1 > \beta^2 \sum_{j=1}^{p} \sigma_{\hat{\Gamma}_j}^{-4}\sigma_{\hat{\gamma}_j}^{4}\kappa_j$ and omitting the term $8\mu_2 v_{12}/(v_2\beta) \sum_{j=1}^{p} \sigma_{\hat{\Gamma}_j}^{-6}\sigma_{\hat{\gamma}_j}^{6}(\kappa_j + 1)$. Therefore, a sufficient condition of $\Delta > 0$ is

$$\sum_{j=1}^{p} \sigma_{\hat{\Gamma}_j}^{-4}\sigma_{\hat{\gamma}_j}^{4}(2\kappa_j + 3)\sum_{j=1}^{p} \sigma_{\hat{\Gamma}_j}^{-4}\sigma_{\hat{\gamma}_j}^{4}(3\kappa_j + 2) > 4\sum_{j=1}^{p} \sigma_{\hat{\Gamma}_j}^{-2}\sigma_{\hat{\gamma}_j}^{2}\kappa_j \sum_{j=1}^{p} \sigma_{\hat{\Gamma}_j}^{-6}\sigma_{\hat{\gamma}_j}^{6}(\kappa_j + 1). \quad \text{(A9)}$$

Since $\hat{\gamma}_j$ and $\hat{\Gamma}_j$ in GWAS are estimated from the marginal linear models, we can further express the ratio of their variances as $\sigma_{\hat{\Gamma}_j}^{-2}\sigma_{\hat{\gamma}_j}^{2} = n_Y \text{Var}(X)(1 - h_{X,j})/\{n_x \text{Var}(Y)(1 - h_{Y,j})\}$, where $h_{X,j}$ and $h_{Y,j}$ denote the variances of $X$ and $Y$ explained by $G_j$, respectively. Let $c_j^2 = (1 - h_{X,j})/(1 - h_{Y,j})$. Then, the inequality (A9) can be simplified as

$$\sum_{j=1}^{p} c_j^4(2\kappa_j + 3)\sum_{j=1}^{p} c_j^4(3\kappa_j + 2) > 4\sum_{j=1}^{p} c_j^2\kappa_j \sum_{j=1}^{p} c_j^6(\kappa_j + 1).$$

Let

$$\Delta_{1j} = c_j^2 - c_j^4 = \left(\frac{1 - h_{X,j}}{1 - h_{Y,j}}\right)\left(\frac{h_{X,j} - h_{Y,j}}{1 - h_{Y,j}}\right),$$

$$\Delta_{2j} = c_j^6 - c_j^4 = -\left(\frac{1 - h_{X,j}}{1 - h_{Y,j}}\right)^2 \left(\frac{h_{X,j} - h_{Y,j}}{1 - h_{Y,j}}\right).$$

Then, we have

$$\sum_{j=1}^{p} c_j^2 \kappa_j \sum_{j=1}^{p} c_j^6 (\kappa_j + 1) = \sum_{j=1}^{p} (c_j^4 + \Delta_{1j})\kappa_j \sum_{j=1}^{p} (c_j^4 + \Delta_{2j})\kappa_j + \sum_{j=1}^{p} (c_j^4 + \Delta_{1j})\kappa_j \sum_{j=1}^{p} (c_j^4 + \Delta_{2j}).$$

Further, we have

$$\sum_{j=1}^{p} c_j^4 (2\kappa_j + 3) \sum_{j=1}^{p} c_j^4 (3\kappa_j + 2) > 6 \sum_{j=1}^{p} c_j^4 \kappa_j \sum_{j=1}^{p} c_j^4 \kappa_j + 13 \sum_{j=1}^{p} c_j^4 \kappa_j \sum_{j=1}^{p} c_j^4$$

Therefore, the inequality (A9) holds when

$$6 \sum_{j=1}^{p} c_j^4 \kappa_j \sum_{j=1}^{p} c_j^4 \kappa_j + 13 \sum_{j=1}^{p} c_j^4 \kappa_j \sum_{j=1}^{p} c_j^4$$
$$> 4 \sum_{j=1}^{p} (c_j^4 + \Delta_{1j})\kappa_j \sum_{j=1}^{p} (c_j^4 + \Delta_{2j})\kappa_j + 4 \sum_{j=1}^{p} (c_j^4 + \Delta_{1j})\kappa_j \sum_{j=1}^{p} (c_j^4 + \Delta_{2j}). \qquad \text{(A10)}$$

Note that, since $\Delta_{1j}\Delta_{2j} < 0$, we have either $\Delta_{1j} > 0$ or $\Delta_{2j} > 0$. Therefore, the inequality (A10) holds when $6c_j^4 > 4(c_j^4 + \Delta_{1j})$ or $6c_j^4 > 4(c_j^4 + \Delta_{2j})$, that is,

$$c_j^4 > 2 \max(\Delta_{1j}, \Delta_{2j}),$$

which holds as long as $\max(h_{X,j}, h_{Y,j}) < 1/3$. Therefore, $\text{Var}(\hat{\beta}_{\text{pIVW}})$ is asymptotically smaller than $\text{Var}(\hat{\beta}_{\text{dIVW}})$ as long as both the proportions of variances of $X$ and $Y$ explained by each IV are less than $1/3$, which is generally true in the genetic context especially when $X$ and $Y$ are some complex traits.

### Web Appendix E: Bootstrapping Fieller's Confidence Interval

Adopting Fieller's method to derive a confidence interval for a ratio, we let $\hat{z}(\beta) = (\tilde{\mu}_1 - \beta\tilde{\mu}_2)^2/(\tilde{v}_1 - 2\beta\tilde{v}_{12} + \beta^2\tilde{v}_2)$, where $\tilde{v}_1 = \hat{\gamma}_j^2\hat{\Gamma}_j^2 - \left(\hat{\Gamma}_j^2 - \sigma_{\hat{\Gamma}_j}^2 - \hat{\tau}^2\right)\left(\hat{\gamma}_j^2 - \sigma_{\hat{\gamma}_j}^2\right)$ and $\tilde{v}_2 = \hat{\mu}_2^2/(2\tilde{\mu}_2 - \hat{\mu}_2)^2 \sum_{j=1}^{p} \sigma_{\hat{\Gamma}_j}^{-4}\left(4\sigma_{\hat{\gamma}_j}^2\hat{\gamma}_j^2 - 2\sigma_{\hat{\gamma}_j}^4\right)$ are the estimated variances of $\tilde{\mu}_1$ and $\tilde{\mu}_2$, respectively, and $\tilde{v}_{12} = 2\tilde{\mu}_2/(2\tilde{\mu}_2 - \hat{\mu}_2)\sum_{j=1}^{p} \sigma_{\hat{\Gamma}_j}^{-4}\sigma_{\hat{\gamma}_j}^2\hat{\gamma}_j\hat{\Gamma}_j$ is the estimated covariance between $\tilde{\mu}_1$ and $\tilde{\mu}_2$. We obtain the $(1 - \alpha)$th quantile $q_{1-\alpha}$ of the distribution of $\hat{z}(\beta)$ via the bootstrap method, where we calculate the $b$th bootstrap statistic $\hat{z}^{(b)}(\hat{\beta}_{\text{pIVW}})$ based on the bootstrap sample $\hat{\gamma}_j^{(b)} \sim N\left(\hat{\gamma}_j, \sigma_{\hat{\gamma}_j}^2\right)$ and $\hat{\Gamma}_j^{(b)} \sim N\left(\hat{\gamma}_j\hat{\beta}_{\text{pIVW}}, \sigma_{\hat{\Gamma}_j}^2 + \hat{\tau}^2\right)$. Then, we solve $\hat{z}(\beta) < q_{1-\alpha}$ for the $100(1-\alpha)\%$ confidence interval of $\beta$. When the IV selection is performed, we construct boostrapping Fieller's confidence interval in a similar way, where only the selected IVs are used to generate the bootstrap samples.

### Web Appendix F: Proof of Theorem 2

The following proof of Theorem 2 takes into account balanced horizontal pleiotropy ($\tau \neq 0$). The situation without horizontal pleiotropy can be considered as a special case with $\tau = 0$.

#### 1 Proof of Theorem 2 (a)

#### 1.1 Bias of dIVW Estimator

When an independent selection dataset $\left\{\gamma_j^*, \sigma_{\hat{\gamma}_j}^*\right\}_{j=1,\ldots p}$ is available, the IVs are included into the analysis when $\left|\gamma_j^*\right| > \delta\sigma_{\hat{\gamma}_j}^*$ with a threshold $\delta$. Then, the dIVW estimator can be written as

$$\hat{\beta}_{\delta,\mathrm{dIVW}} = \frac{\hat{\mu}_{1,\delta}}{\hat{\mu}_{2,\delta}} = \frac{\sum_{j=1}^{p}\sigma_{\hat{\Gamma}_j}^{-2}\hat{\gamma}_j\hat{\Gamma}_j s_j}{\sum_{j=1}^{p}\sigma_{\hat{\Gamma}_j}^{-2}\left(\hat{\gamma}_j^2 - \sigma_{\hat{\gamma}_j}^2\right)s_j},$$

where $s_j = I\left\{\left|\gamma_j^*\right| > \delta\sigma_{\hat{\gamma}_j}^*\right\}$ with the indicative function $I\{\cdot\}$.

Similar to the proof in Section 1.1 of Web Appendix C, the bias of the dIVW estimator is

$$E\left(\hat{\beta}_{\delta,\mathrm{dIVW}} - \beta\right) = \beta\left(\frac{v_{2,\delta}}{\mu_{2,\delta}^2} - \frac{v_{12,\delta}}{\mu_{1,\delta}\mu_{2,\delta}}\right) + o(\xi_\delta) = O(\xi_\delta),$$

where $\mu_{1,\delta} = E\left(\hat{\mu}_{1,\delta}\right) = \beta\sum_{j=1}^{p}\sigma_{\hat{\Gamma}_j}^{-2}\gamma_j^2 q_{\delta,j} = \Theta(\kappa_\delta p_\delta)$, $\mu_{2,\delta} = E\left(\hat{\mu}_{2,\delta}\right) = \sum_{j=1}^{p}\sigma_{\hat{\Gamma}_j}^{-2}\gamma_j^2 q_{\delta,j} = \Theta(\kappa_\delta p_\delta)$, $v_{2,\delta} = \mathrm{Var}\left(\hat{\mu}_{2,\delta}\right) = \sum_{j=1}^{p}\sigma_{\hat{\Gamma}_j}^{-4}\left(4\sigma_{\hat{\gamma}_j}^2\gamma_j^2 + 2\sigma_{\hat{\gamma}_j}^4\right)q_{\delta,j} + \sum_{j=1}^{p}\sigma_{\hat{\Gamma}_j}^{-4}\gamma_j^4 q_{\delta,j}\left(1 - q_{\delta,j}\right) = \Theta(\kappa_\delta p_\delta + p_\delta) + \Theta(\varphi^2 p_\delta)$ and $v_{12,\delta} = \mathrm{Cov}\left(\hat{\mu}_{1,\delta}, \hat{\mu}_{2,\delta}\right) = 2\beta\sum_{j=1}^{p}\sigma_{\hat{\Gamma}_j}^{-4}\sigma_{\hat{\gamma}_j}^2\gamma_j^2 q_{\delta,j} + \beta\sum_{j=1}^{p}\sigma_{\hat{\Gamma}_j}^{-4}\gamma_j^4 q_{\delta,j}\left(1 - q_{\delta,j}\right) = \Theta(\kappa_\delta p_\delta) + \Theta(\varphi^2 p_\delta)$ under Assumptions 2 and 3. As the effective sample size $\eta_\delta = \kappa_\delta\sqrt{p_\delta}/\max(1,\varphi) \to \infty$, $\xi_\delta = 1/\kappa_\delta p_\delta + \max(1,\varphi^2)/\kappa_\delta^2 p_\delta$ converges to zero.

#### 1.2 Bias of pIVW Estimator

The proposed pIVW estimator $\hat{\beta}_{\delta,\mathrm{pIVW}}$ can be written as

$$\hat{\beta}_{\delta,\mathrm{pIVW}} = \frac{\hat{\mu}_{1,\delta}}{\tilde{\mu}_{2,\delta}} + \frac{\hat{v}_{12,\delta}}{\hat{v}_{2,\delta}}\left(1 - \frac{\hat{\mu}_{2,\delta}}{\tilde{\mu}_{2,\delta}}\right),$$

where $\hat{v}_{2,\delta} = \sum_{j=1}^{p}\sigma_{\hat{\Gamma}_j}^{-4}\left\{4\left(\hat{\gamma}_j^2 - \sigma_{\hat{\gamma}_j}^2\right)\sigma_{\hat{\gamma}_j}^2 + 2\sigma_{\hat{\gamma}_j}^4\right\}s_j$, $\hat{v}_{12,\delta} = 2\sum_{j=1}^{p}\sigma_{\hat{\Gamma}_j}^{-4}\sigma_{\hat{\gamma}_j}^2\hat{\Gamma}_j\hat{\gamma}_j s_j$ and $\tilde{\mu}_{2,\delta} = \hat{\mu}_{2,\delta}\left(1 + \sqrt{1 + 4\lambda\hat{v}_{2,\delta}/\hat{\mu}_{2,\delta}^2}\right)/2$. As in Section 1.2 of Web Appendix C, we have

$$E\left(\frac{\hat{\mu}_{1,\delta}}{\tilde{\mu}_{2,\delta}} - \frac{\mu_{1,\delta}}{\mu_{2,\delta}}\right) = -\frac{\beta\lambda v_{2,\delta}^*}{\mu_{2,\delta}^2} + \beta\left(\frac{v_{2,\delta}}{\mu_{2,\delta}^2} - \frac{v_{12,\delta}}{\mu_{1,\delta}\mu_{2,\delta}}\right) + o(\xi_\delta),$$

$$E\left(\frac{\hat{v}_{12,\delta}}{\hat{v}_{2,\delta}}\right)\left(1 - \frac{\hat{\mu}_{2,\delta}}{\tilde{\mu}_{2,\delta}}\right) = \frac{\lambda v_{12,\delta}^*}{\mu_{2,\delta}^2} + o(\xi_\delta),$$

where $v_{2,\delta}^* = E(\hat{v}_{2,\delta}) = \sum_{j=1}^{p} \sigma_{\hat{\Gamma}_j}^{-4}\left(4\sigma_{\hat{\gamma}_j}^2\gamma_j^2 + 2\sigma_{\hat{\gamma}_j}^4\right) q_{\delta,j} = \Theta(\kappa_\delta p_\delta + p_\delta)$ and $v_{12,\delta}^* = E(\hat{v}_{12,\delta}) = 2\beta \sum_{j=1}^{p} \sigma_{\hat{\Gamma}_j}^{-4}\sigma_{\hat{\gamma}_j}^2\gamma_j^2 q_{\delta,j} = \Theta(\kappa_\delta p_\delta)$. Then, the bias of the pIVW estimator is

$$E(\hat{\beta}_{\delta,\mathrm{pIVW}} - \beta) = E\left(\frac{\hat{\mu}_{1,\delta}}{\tilde{\mu}_{2,\delta}} - \frac{\mu_{1,\delta}}{\mu_{2,\delta}}\right) + E\left(\frac{\hat{v}_{12,\delta}}{\hat{v}_{2,\delta}}\right)\left(1 - \frac{\hat{\mu}_{2,\delta}}{\tilde{\mu}_{2,\delta}}\right)$$
$$= (1-\lambda)\beta\left(\frac{v_{2,\delta}}{\mu_{2,\delta}^2} - \frac{v_{12,\delta}}{\mu_{1,\delta}\mu_{2,\delta}}\right) + o(\xi_\delta).$$

### 2 Proof of Theorem 2 (b)

Similar to the proof in Section 2 of Web Appendix C, we have

$$E(\hat{\beta}_{\delta,\mathrm{dIVW}} - \beta)^2$$
$$= \beta^2\left\{\frac{v_{1,\delta}}{\mu_{1,\delta}^2} + \frac{v_{2,\delta}}{\mu_{2,\delta}^2} - \frac{2v_{12,\delta}}{\mu_{1,\delta}\mu_{2,\delta}} - 2E(\hat{x}_{1,\delta}^2\hat{x}_{2,\delta}) + 4E(\hat{x}_{1,\delta}\hat{x}_{2,\delta}^2) - 2E(\hat{x}_{2,\delta}^3)\right.$$
$$\left. + 3E(\hat{x}_{1,\delta}^2\hat{x}_{2,\delta}^2) - 6E(\hat{x}_{1,\delta}\hat{x}_{2,\delta}^3) + 3E(\hat{x}_{2,\delta}^4)\right\} + o(\xi_\delta^2)$$
$$= O(\xi_\delta). \qquad \text{(A11)}$$

where $v_{1,\delta} = \mathrm{Var}(\hat{\mu}_{1,\delta}) = \sum_{j=1}^{p} \sigma_{\hat{\Gamma}_j}^{-4}\left\{\left(\sigma_{\hat{\gamma}_j}^2\beta^2 + \sigma_{\hat{\Gamma}_j}^2 + \tau^2\right)\gamma_j^2 + \left(\sigma_{\hat{\Gamma}_j}^2 + \tau^2\right)\sigma_{\hat{\gamma}_j}^2\right\} q_{\delta,j} + \beta^2 \sum_{j=1}^{p} \sigma_{\hat{\Gamma}_j}^{-4}\gamma_j^4 q_{\delta,j}(1 - q_{\delta,j}) = \Theta(\kappa_\delta p_\delta + p_\delta) + \Theta(\varphi^2 p_\delta)$. Therefore, the variance of the dIVW estimator is

$$\mathrm{Var}(\hat{\beta}_{\delta,\mathrm{dIVW}}) = E(\hat{\beta}_{\delta,\mathrm{dIVW}} - \beta)^2 - E^2(\hat{\beta}_{\delta,\mathrm{dIVW}} - \beta) = O(\xi_\delta).$$

The difference in variance between the dIVW estimator and the pIVW estimator is

$$\mathrm{Var}(\hat{\beta}_{\delta,\mathrm{dIVW}}) - \mathrm{Var}(\hat{\beta}_{\delta,\mathrm{pIVW}})$$
$$= E(\hat{\beta}_{\delta,\mathrm{dIVW}} - \beta)^2 - E\left(\hat{\beta}_{\delta,\mathrm{pIVW}} - \frac{\mu_{1,\delta}}{m_{2,\delta}}\right)^2 + E^2\left(\hat{\beta}_{\delta,\mathrm{pIVW}} - \frac{\mu_{1,\delta}}{m_{2,\delta}}\right) - E^2(\hat{\beta}_{\delta,\mathrm{dIVW}} - \beta),$$

where $m_{2,\delta} = E(\tilde{\mu}_{2,\delta})$. Similar to that in Section 2 of Web Appendix C, we have

$$E^2\left(\hat{\beta}_{\delta,\mathrm{pIVW}} - \frac{\mu_{1,\delta}}{m_{2,\delta}}\right) = \left\{E(\hat{\beta}_{\delta,\mathrm{pIVW}} - \beta) + \left(\beta - \frac{\mu_{1,\delta}}{m_{2,\delta}}\right)\right\}^2$$
$$= \beta^2\left\{(1-\lambda)\left(\frac{v_{2,\delta}^*}{\mu_{2,\delta}^2} - \frac{v_{12,\delta}^*}{\mu_{1,\delta}\mu_{2,\delta}}\right) + \frac{\lambda v_{2,\delta}^*}{\mu_{2,\delta}^2}\right\}^2 + o(\xi_\delta^2),$$

and therefore

$$E^2\left(\hat{\beta}_{\delta,\mathrm{pIVW}} - \frac{\mu_{1,\delta}}{m_{2,\delta}}\right) - E^2(\hat{\beta}_{\delta,\mathrm{dIVW}} - \beta) = \frac{2\lambda\beta v_{2,\delta}^* v_{12,\delta}^*}{\mu_{2,\delta}^4} + \frac{v_{12,\delta}^{*2}}{\mu_{2,\delta}^4}(\lambda^2 - 2\lambda) + o(\xi_\delta^2)$$
$$\text{(A12)}$$

Next, we calculate $E(\hat{\beta}_{\delta,\mathrm{dIVW}} - \beta)^2 - E\left(\hat{\beta}_{\delta,\mathrm{pIVW}} - \frac{\mu_{1,\delta}}{m_{2,\delta}}\right)^2$. By Taylor series expansion,

$$E\left(\hat{\beta}_{\delta,\mathrm{pIVW}}-\frac{\mu_{1,\delta}}{m_{2,\delta}}\right)^2$$

$$\begin{aligned}
&= \left(\frac{\mu_{1,\delta}}{m_{2,\delta}}\right)^2\left(\frac{v_{1,\delta}}{\mu_{1,\delta}^2}+\frac{w_\delta^2 v_{2,\delta}}{m_{2,\delta}^2}-\frac{2w_\delta v_{12,\delta}}{\mu_{1,\delta}m_{2,\delta}}\right)\\
&+\beta^2\{-2E(\hat{x}_{1,\delta}^2\hat{x}_{2,\delta})+4E(\hat{x}_{1,\delta}\hat{x}_{2,\delta}^2)-2E(\hat{x}_{2,\delta}^3)+3E(\hat{x}_{1,\delta}^2\hat{x}_{2,\delta}^2)\\
&-6E(\hat{x}_{1,\delta}\hat{x}_{2,\delta}^3)+3E(\hat{x}_{2,\delta}^4)\}+\frac{\lambda^2 v_{12,\delta}^{*2}}{\mu_{2,\delta}^4}+\frac{6\lambda\beta v_{12,\delta}^*}{\mu_{2,\delta}^2}\left(\frac{v_{2,\delta}^*}{\mu_{2,\delta}^2}-\frac{v_{12,\delta}^*}{\mu_{1,\delta}\mu_{2,\delta}}\right)\\
&-16\lambda\beta\left(\frac{v_{12,\delta}^*}{v_{2,\delta}^*}-\beta\right)\frac{\sum_{j=1}^p\sigma_{\hat{\Gamma}_j}^{-6}\sigma_{\hat{\gamma}_j}^6\left(\gamma_j^2\sigma_{\hat{\gamma}_j}^{-2}+1\right)q_j}{\mu_{2,\delta}^3}+o(\xi_\delta^2), \qquad \text{(A13)}
\end{aligned}$$

where $w_\delta=\mu_{2,\delta}^*/(2\mu_{2,\delta}^*-\mu_{2,\delta})$ with $\mu_{2,\delta}^*=\mu_{2,\delta}\left(1+\sqrt{1+4\lambda v_{2,\delta}^*/\mu_{2,\delta}^2}\right)/2$. From Equations (A11) and (A13), we have

$$E(\hat{\beta}_{\delta,\mathrm{dIVW}}-\beta)^2-E\left(\hat{\beta}_{\delta,\mathrm{pIVW}}-\frac{\mu_{1,\delta}}{m_{2,\delta}}\right)^2$$

$$\begin{aligned}
&=\frac{2\lambda v_{1,\delta}^* v_{2,\delta}^*}{\mu_{2,\delta}^4}+\frac{6\lambda\beta^2 v_{2,\delta}^{*2}}{\mu_{2,\delta}^4}-\frac{14\lambda\beta v_{2,\delta}^* v_{12,\delta}^*}{\mu_{2,\delta}^4}+\frac{(6\lambda-\lambda^2)v_{12,\delta}^{*2}}{\mu_{2,\delta}^4}\\
&+16\lambda\beta\left(\frac{v_{12,\delta}^*}{v_{2,\delta}^*}-\beta\right)\frac{\sum_{j=1}^p\sigma_{\hat{\Gamma}_j}^{-6}\sigma_{\hat{\gamma}_j}^6\left(\gamma_j^2\sigma_{\hat{\gamma}_j}^{-2}+1\right)q_j}{\mu_{2,\delta}^3}+o(\xi_\delta^2), \qquad \text{(A14)}
\end{aligned}$$

where $v_{1,\delta}^*=\sum_{j=1}^p\sigma_{\hat{\Gamma}_j}^{-4}\left\{\left(\sigma_{\hat{\gamma}_j}^2\beta^2+\sigma_{\hat{\Gamma}_j}^2+\tau^2\right)\gamma_j^2+\left(\sigma_{\hat{\Gamma}_j}^2+\tau^2\right)\sigma_{\hat{\gamma}_j}^2\right\}q_{\delta,j}=\Theta(\kappa_\delta p_\delta+p_\delta)$. Combining Equations (A12) and (A14), the difference in the variance between the dIVW estimator and the pIVW estimator is

$$\begin{aligned}
&\mathrm{Var}(\hat{\beta}_{\delta,\mathrm{dIVW}})-\mathrm{Var}(\hat{\beta}_{\delta,\mathrm{pIVW}})\\
&=\frac{2\lambda v_{1,\delta}^* v_{2,\delta}^*}{\mu_{2,\delta}^4}+\frac{6\lambda\beta^2 v_{2,\delta}^{*2}}{\mu_{2,\delta}^4}-\frac{12\lambda\beta v_{2,\delta}^* v_{12,\delta}^*}{\mu_{2,\delta}^4}+\frac{4\lambda v_{12,\delta}^{*2}}{\mu_{2,\delta}^4}\\
&+16\lambda\beta\left(\frac{v_{12,\delta}^*}{v_{2,\delta}^*}-\beta\right)\frac{\sum_{j=1}^p\sigma_{\hat{\Gamma}_j}^{-6}\sigma_{\hat{\gamma}_j}^6\left(\gamma_j^2\sigma_{\hat{\gamma}_j}^{-2}+1\right)q_j}{\mu_{2,\delta}^3}+o(\xi_\delta^2)\\
&=\frac{2\lambda\beta^2}{\mu_{2,\delta}^4}\Delta_\delta+o(\xi_\delta^2),
\end{aligned}$$

where

$$\begin{aligned}
\Delta_\delta=\frac{3(\mu_{1,\delta}v_{2,\delta}^*-\mu_{2,\delta}v_{12,\delta}^*)^2}{\mu_{2,\delta}^2\beta^2}+\frac{v_{1,\delta}^*v_{2,\delta}^*-v_{12,\delta}^{*2}}{\beta^2}\\
+8\mu_{2,\delta}\left(\frac{v_{12,\delta}^*}{v_{2,\delta}^*\beta}-1\right)\sum_{j=1}^p\sigma_{\hat{\Gamma}_j}^{-6}\sigma_{\hat{\gamma}_j}^6\left(\gamma_j^2\sigma_{\hat{\gamma}_j}^{-2}+1\right)q_j.
\end{aligned}$$

Similar to the proof in Web Appendix D, it can be shown that $\Delta_\delta>0$ when both the proportions of variances of $X$ and $Y$ explained by each IV are less than $1/3$.

**3 Proof of Theorem 2 (c)**

To prove

$$V_\delta^{-\frac{1}{2}}(\hat{\beta}_{\delta,\mathrm{pIVW}} - \beta) = \left(\frac{\mu_{2,\delta}}{\sqrt{v_\delta}}\right)\frac{\hat{\mu}_{1,\delta} - \beta\hat{\mu}_{2,\delta}}{\tilde{\mu}_{2,\delta}} + \left(\frac{\mu_{2,\delta}}{\sqrt{v_\delta}}\right)\left(\frac{\hat{\mu}_{2,\delta}}{\tilde{\mu}_{2,\delta}} - 1\right)\left(\beta - \frac{\hat{v}_{12,\delta}}{\hat{v}_{2,\delta}}\right) \xrightarrow{d} N(0,1),$$

where $V_\delta = \mu_{2,\delta}^{-2}\sum_{j=1}^{p}\left\{\sigma_{\hat{\Gamma}_j}^{-2}\left(\gamma_j^2 + \sigma_{\hat{\gamma}_j}^2\right)\left(1 + \tau^2\sigma_{\hat{\Gamma}_j}^{-2}\right) + \beta^2\sigma_{\hat{\gamma}_j}^2\sigma_{\hat{\Gamma}_j}^{-4}\left(\gamma_j^2 + 2\sigma_{\hat{\gamma}_j}^2\right)\right\}q_{\delta,j}$ and $v_\delta = \mu_{2,\delta}^2 V_\delta = \Theta(\kappa_\delta p_\delta + p_\delta)$, it suffices to prove $\left(\frac{\mu_{2,\delta}}{\sqrt{v_\delta}}\right)\frac{\hat{\mu}_{1,\delta} - \beta\hat{\mu}_{2,\delta}}{\tilde{\mu}_{2,\delta}} \xrightarrow{d} N(0,1)$ and $\left(\frac{\mu_{2,\delta}}{\sqrt{v_\delta}}\right)\left(\frac{\hat{\mu}_{2,\delta}}{\tilde{\mu}_{2,\delta}} - 1\right)\left(\beta - \frac{\hat{v}_{12,\delta}}{\hat{v}_{2,\delta}}\right) \xrightarrow{p} 0$. As the proof in Section 3 of Web Appendix C, we first prove $\tilde{\mu}_{2,\delta}/\mu_{2,\delta} \xrightarrow{p} 1$ as $\eta_\delta \to \infty$. Then, we prove $\frac{\hat{\mu}_{1,\delta} - \beta\hat{\mu}_{2,\delta}}{\sqrt{v_\delta}} \xrightarrow{d} N(0,1)$ as $p \to \infty$ by the Lindeberg Central Limit Theorem provided that $\max_j \gamma_j^2\sigma_{\hat{\gamma}_j}^{-2}q_{\delta,j}/(\kappa_\delta p_\delta + p_\delta) \to 0$. Lastly, we prove $\left(\frac{\mu_{2,\delta}}{\sqrt{v_\delta}}\right)\left(\frac{\hat{\mu}_{2,\delta}}{\tilde{\mu}_{2,\delta}} - 1\right)\left(\beta - \frac{\hat{v}_{12,\delta}}{\hat{v}_{2,\delta}}\right) \xrightarrow{p} 0$ by showing $\left(\frac{\mu_{2,\delta}}{\sqrt{v_\delta}}\right)\left(\frac{\hat{\mu}_{2,\delta}}{\tilde{\mu}_{2,\delta}} - 1\right) = \left(\frac{\lambda v_{2,\delta}^*}{\sqrt{v_\delta}\mu_{2,\delta}}\right)\frac{\mu_{2,\delta}^2}{\hat{\mu}_{2,\delta}^2}\frac{\hat{v}_{2,\delta}}{v_{2,\delta}^*}\frac{1}{r_{\lambda,\delta}^2} \xrightarrow{p} 0$ and $\beta - \frac{\hat{v}_{12,\delta}}{\hat{v}_{2,\delta}} = O_p(1)$ as $\eta_\delta \to \infty$. Further, the normality of $\hat{V}_\delta^{-\frac{1}{2}}(\hat{\beta}_{\delta,\mathrm{pIVW}} - \beta)$ with the plug-in estimator $\hat{V}_\delta$ of $V_\delta$ follows from $\tilde{\mu}_{2,\delta}/\mu_{2,\delta} \xrightarrow{p} 1$ and $\hat{v}_\delta/v_\delta \xrightarrow{p} 1$ as $\eta_\delta \to \infty$. We omit the details here.

### Web Appendix G: Simulation with Individual-Level Data

First, we randomly generate the individual-level data for the genetic variants from $G_j \sim Bin(2, \mathrm{MAF}_j)$ where the minor allele frequencies $\mathrm{MAF}_j \sim U(0.1, 0.5)$. Then, we simulate the individual-level data for the exposure $X$ and the outcome $Y$ based on the linear structural models (1) and (7), where we consider similar settings of model parameters as in Section 4.1. Specifically, we set $\beta = 0.5$, and $U, \epsilon_X$ and $\epsilon_Y$ are generated from $N(0, 2)$ independently. The IV effects $\gamma_j$ and the balanced horizontal pleiotropy $\alpha_j$ are generated in the same ways as in Section 4.1. We set the sample sizes of the exposure dataset $n_X = 100{,}000$ and the outcome dataset $n_Y = 2n_X$. To consider the IV selection, we generate an independent dataset based on model (1) with the sample size $n_X^* = 2n_X$. Lastly, we obtain the summary-level data $\left\{\hat{\gamma}_j^*, \sigma_{\hat{\gamma}_j}^*\right\}_{j=1}^{1000}$, $\left\{\hat{\gamma}_j, \sigma_{\hat{\gamma}_j}\right\}_{j=1}^{1000}$ and $\left\{\hat{\Gamma}_j, \sigma_{\hat{\Gamma}_j}\right\}_{j=1}^{1000}$ by estimating the marginal effects and their standard errors in the corresponding linear regressions, which are based on three independent datasets for the selection, the exposure, and the outcome data respectively. The simulation results are provided in Web Tables 6-9.

### Web Appendix H: Estimation of the Effective Sample Size

When no IV selection is performed, we estimate the effective sample size $\eta$ by

$$\hat{\eta} = \hat{\kappa}\sqrt{p},$$

where the estimated IV strength $\hat{\kappa} = p^{-1}\sum_{j=1}^{p}\left(\hat{\gamma}_j^2 - \sigma_{\hat{\gamma}_j}^{-2}\right)\sigma_{\hat{\gamma}_j}^2$.

When IV selection is performed, we estimate the effective sample size $\eta_\delta$ by

$$\hat{\eta}_\delta = \frac{\hat{\kappa}_\delta\sqrt{\hat{p}_\delta}}{\max(1, \hat{\varphi})},$$

where $\hat{p}_\delta$ is the number of selected IVs within the set $S_\delta$, the estimated IV strength $\hat{\kappa}_\delta = \hat{p}_\delta^{-1}\sum_{j\in S_\delta}\left(\hat{\gamma}_j^2 - \sigma_{\hat{\gamma}_j}^{-2}\right)\sigma_{\hat{\gamma}_j}^2$ , and $\hat{\varphi} = \sqrt{\hat{p}_\delta^{-1}\sum_{j=1}^{p}\left(\hat{\gamma}_j^4\sigma_{\hat{\gamma}_j}^{-4} - 6\hat{\gamma}_j^2\sigma_{\hat{\gamma}_j}^{-2} + 3\right)\hat{q}_{\delta,j}\left(1 - \hat{q}_{\delta,j}\right)}$ is the estimate of $\varphi$, where $\hat{q}_{\delta,j} = \Phi\left(\hat{\gamma}_j^*/\sigma_{\hat{\gamma}_j}^* - \delta\right) + \Phi\left(-\hat{\gamma}_j^*/\sigma_{\hat{\gamma}_j}^* - \delta\right)$ and $\Phi(\cdot)$ is the cumulative distribution function for standard normal distribution.

### Web Tables

#### Web Table 1

The pIVW estimator with various penalty parameter $\lambda$. The true causal effect $\beta = 0.5$ and no horizontal pleiotropy exists ($\tau = 0$). The IV selection threshold $\delta = \sqrt{2 \log p}$. The simulation is based on 10,000 replicates. Bias (%): bias divided by $\beta$; SE: empirical standard error; CP (%): coverage probability of the 95% confidence interval.

| $\eta_\delta$ | $\lambda$ | Bias | SE | CP |
|---|---|---|---|---|
| 6.76 | 0 | 3.2 | 0.144 | 95.7 |
| | 0.5 | 1.5 | 0.140 | 95.5 |
| | 1 | -0.1 | 0.136 | 95.3 |
| | 1.5 | -1.4 | 0.133 | 95.2 |
| | 2 | -2.6 | 0.131 | 95.2 |
| | 2.5 | -3.7 | 0.129 | 95.0 |
| 10.26 | 0 | 1.4 | 0.090 | 95.1 |
| | 0.5 | 0.7 | 0.089 | 95.1 |
| | 1 | 0.1 | 0.088 | 95.1 |
| | 1.5 | -0.6 | 0.088 | 95.0 |
| | 2 | -1.2 | 0.087 | 95.1 |
| | 2.5 | -1.8 | 0.086 | 95.1 |
| 17.84 | 0 | 0.7 | 0.056 | 94.7 |
| | 0.5 | 0.4 | 0.055 | 94.7 |
| | 1 | 0.1 | 0.055 | 94.7 |
| | 1.5 | -0.1 | 0.055 | 94.7 |
| | 2 | -0.4 | 0.055 | 94.7 |
| | 2.5 | -0.6 | 0.055 | 94.7 |

### Web Table 2

The pIVW estimator with various penalty parameter $\lambda$. The true causal effect $\beta = 0.5$ and balanced horizontal pleiotropy exists $\tau = 0.01$. No IV selection is conducted. The simulation is based on 10,000 replicates. Bias (%): bias divided by $\beta$; SE: empirical standard error; CP (%): coverage probability of the 95% confidence interval.

| $\eta$ | $\lambda$ | Bias | SE | CP |
|---|---|---|---|---|
| 4.33 | 0 | 28.0 | 2.441 | 94.9 |
| | 0.5 | 5.1 | 0.444 | 94.7 |
| | 1 | -2.2 | 0.389 | 94.4 |
| | 1.5 | -7.5 | 0.357 | 94.2 |
| | 2 | -11.6 | 0.335 | 93.9 |
| | 2.5 | -14.9 | 0.318 | 93.3 |
| 9.52 | 0 | 2.9 | 0.194 | 95.0 |
| | 0.5 | 1.2 | 0.189 | 94.8 |
| | 1 | -0.3 | 0.185 | 94.8 |
| | 1.5 | -1.7 | 0.181 | 94.6 |
| | 2 | -3.0 | 0.178 | 94.4 |
| | 2.5 | -4.3 | 0.175 | 94.3 |
| 21.85 | 0 | 0.5 | 0.091 | 94.7 |
| | 0.5 | 0.1 | 0.091 | 94.7 |
| | 1 | -0.1 | 0.090 | 94.7 |
| | 1.5 | -0.6 | 0.090 | 94.6 |
| | 2 | -0.9 | 0.090 | 94.6 |
| | 2.5 | -1.2 | 0.089 | 94.5 |

### Web Table 3

The pIVW estimator with various penalty parameter $\lambda$. The true causal effect $\beta = 0.5$ and balanced horizontal pleiotropy exists ($\tau = 0.01$). The IV selection threshold $\delta = \sqrt{2\log p}$. The simulation is based on 10,000 replicates. Bias (%): bias divided by $\beta$; SE: empirical standard error; CP (%): coverage probability of the 95% confidence interval.

| $\eta_\delta$ | $\lambda$ | Bias | SE | CP |
|---|---|---|---|---|
| 6.76 | 0 | 3.9 | 0.198 | 95.1 |
| | 0.5 | 2.2 | 0.193 | 95.0 |
| | 1 | 0.7 | 0.189 | 94.9 |
| | 1.5 | -0.7 | 0.186 | 94.9 |
| | 2 | -1.9 | 0.183 | 94.9 |
| | 2.5 | -3.0 | 0.180 | 94.9 |
| 10.26 | 0 | 1.2 | 0.125 | 94.9 |
| | 0.5 | 0.5 | 0.124 | 94.8 |
| | 1 | -0.2 | 0.123 | 94.8 |
| | 1.5 | -0.8 | 0.122 | 94.7 |
| | 2 | -1.4 | 0.121 | 94.7 |
| | 2.5 | -2.0 | 0.120 | 94.6 |
| 17.84 | 0 | 0.4 | 0.078 | 94.9 |
| | 0.5 | 0.1 | 0.078 | 94.9 |
| | 1 | -0.2 | 0.078 | 94.9 |
| | 1.5 | -0.4 | 0.078 | 94.8 |
| | 2 | -0.7 | 0.077 | 94.8 |
| | 2.5 | -0.9 | 0.077 | 94.8 |

### Web Table 4

Comparison of the pIVW estimator ($\lambda_{opt}$ = 1) with other competing MR methods. The true causal effect $\beta = 0.5$ and balanced horizontal pleiotropy exists ($\tau = 0.01$). No IV selection is conducted. The simulation is based on 10,000 replicates. Bias (%): bias divided by $\beta$; SE: empirical standard error; CP (%): coverage probability of the 95% confidence interval.

| $\eta$ | Method | Bias | SE | CP |
|---|---|---|---|---|
| 4.33 | IVW | -88.0 | 0.040 | 0.0 |
| | MR-Egger | -80.1 | 0.065 | 0.0 |
| | MR-Median | -81.7 | 0.057 | 0.0 |
| | MR-RAPS | 33.1 | 2.471 | 96.3 |
| | dIVW | 28.0 | 2.441 | 96.4 |
| | pIVW | -2.2 | 0.389 | 94.4 |
| 9.52 | IVW | -76.9 | 0.038 | 0.0 |
| | MR-Egger | -63.5 | 0.059 | 0.0 |
| | MR-Median | -66.0 | 0.056 | 0.0 |
| | MR-RAPS | 2.9 | 0.195 | 95.7 |
| | dIVW | 2.9 | 0.194 | 95.9 |
| | pIVW | -0.3 | 0.185 | 94.8 |
| 21.85 | IVW | -59.2 | 0.034 | 0.0 |
| | MR-Egger | -42.0 | 0.051 | 1.6 |
| | MR-Median | -46.8 | 0.050 | 0.2 |
| | MR-RAPS | 0.5 | 0.093 | 94.9 |
| | dIVW | 0.5 | 0.091 | 94.9 |
| | pIVW | -0.2 | 0.090 | 94.7 |

### Web Table 5

Comparison of the pIVW estimator ($\lambda_{opt}$ = 1) with other competing MR methods. The true causal effect $\beta = 0.5$ and balanced horizontal pleiotropy exists ($\tau = 0.01$). The IV selection threshold $\delta = \sqrt{2 \log p}$. The simulation is based on 10,000 replicates. Bias (%): bias divided by $\beta$; SE: empirical standard error; CP (%): coverage probability of the 95% confidence interval.

| $\eta_\delta$ | Method | Bias | SE | CP |
|---|---|---|---|---|
| 6.76 | IVW | -6.6 | 0.172 | 90.6 |
| | MR-Egger | -42.7 | 0.534 | 88.1 |
| | MR-Median | -11.3 | 0.199 | 88.3 |
| | MR-RAPS | 2.8 | 0.193 | 92.8 |
| | dIVW | 3.9 | 0.198 | 95.8 |
| | pIVW | 0.7 | 0.189 | 94.9 |
| 10.26 | IVW | -8.1 | 0.111 | 91.0 |
| | MR-Egger | -44.2 | 0.289 | 83.7 |
| | MR-Median | -12.6 | 0.137 | 86.3 |
| | MR-RAPS | 0.9 | 0.124 | 93.1 |
| | dIVW | 1.2 | 0.125 | 95.1 |
| | pIVW | -0.2 | 0.123 | 94.8 |
| 17.84 | IVW | -8.1 | 0.070 | 89.4 |
| | MR-Egger | -48.8 | 0.183 | 70.8 |
| | MR-Median | -12.8 | 0.089 | 82.3 |
| | MR-RAPS | 0.4 | 0.078 | 93.7 |
| | dIVW | 0.4 | 0.078 | 95.0 |
| | pIVW | -0.2 | 0.078 | 94.9 |

### Web Table 6

The simulation with individual-level data. Comparison of the pIVW estimator ($\lambda_{opt}$ = 1) with other competing MR methods. The true causal effect $\beta = 0.5$. No horizontal pleiotropy exists ($\tau = 0$). No IV selection is conducted. The simulation is based on 10,000 replicates. Bias (%): bias divided by $\beta$; SE: empirical standard error; CP (%): coverage probability of the 95% confidence interval.

| $\eta$ | Method | Bias | SE | CP |
|---|---|---|---|---|
| 4.33 | IVW | -87.9 | 0.028 | 0.0 |
| | MR-Egger | -80.0 | 0.045 | 0.0 |
| | MR-Median | -83.0 | 0.041 | 0.0 |
| | MR-RAPS | 8.4 | 0.694 | 94.1 |
| | dIVW | 18.0 | 2.535 | 95.2 |
| | pIVW | -1.9 | 0.298 | 94.2 |
| 9.52 | IVW | -76.8 | 0.026 | 0.0 |
| | MR-Egger | -63.3 | 0.041 | 0.0 |
| | MR-Median | -67.5 | 0.040 | 0.0 |
| | MR-RAPS | 0.9 | 0.118 | 95.2 |
| | dIVW | 3.1 | 0.148 | 95.7 |
| | pIVW | -0.1 | 0.139 | 95.0 |
| 21.85 | IVW | -59.0 | 0.023 | 0.0 |
| | MR-Egger | -41.8 | 0.035 | 0.0 |
| | MR-Median | -46.6 | 0.035 | 0.0 |
| | MR-RAPS | 0.1 | 0.058 | 95.2 |
| | dIVW | 0.6 | 0.067 | 95.5 |
| | pIVW | -0.1 | 0.066 | 95.3 |

### Web Table 7

The simulation with individual-level data. Comparison of the pIVW estimator ($\lambda_{opt}$ = 1) with other competing MR methods. The true causal effect $\beta = 0.5$. No horizontal pleiotropy exists ($\tau = 0$). The IV selection threshold $\delta = \sqrt{2 \log p}$. The simulation is based on 10,000 replicates. Bias (%): bias divided by $\beta$; SE: empirical standard error; CP (%): coverage probability of the 95% confidence interval.

| $\eta_\delta$ | Method | Bias | SE | CP |
|---|---|---|---|---|
| 6.76 | IVW | -7.4 | 0.120 | 93.7 |
| | MR-Egger | -41.6 | 0.384 | 89.0 |
| | MR-Median | -11.7 | 0.139 | 95.0 |
| | MR-RAPS | 1.4 | 0.135 | 95.9 |
| | dIVW | 3.1 | 0.142 | 96.1 |
| | pIVW | -0.1 | 0.134 | 95.4 |
| 10.26 | IVW | -8.2 | 0.078 | 91.3 |
| | MR-Egger | -43.1 | 0.204 | 79.4 |
| | MR-Median | -12.3 | 0.095 | 93.1 |
| | MR-RAPS | 0.2 | 0.087 | 95.4 |
| | dIVW | 0.9 | 0.090 | 95.4 |
| | pIVW | -0.4 | 0.088 | 95.0 |
| 17.84 | IVW | -7.9 | 0.049 | 86.9 |
| | MR-Egger | -48.5 | 0.129 | 51.0 |
| | MR-Median | -12.3 | 0.061 | 88.7 |
| | MR-RAPS | 0.5 | 0.054 | 95.4 |
| | dIVW | 0.7 | 0.055 | 95.4 |
| | pIVW | 0.2 | 0.055 | 94.9 |

### Web Table 8

The simulation with individual-level data. Comparison of the pIVW estimator ($\lambda_{opt}$ = 1) with other competing MR methods. The true causal effect $\beta = 0.5$ and balanced horizontal pleiotropy exists ($\tau = 0.01$). No IV selection is conducted. The simulation is based on 10,000 replicates. Bias (%): bias divided by $\beta$; SE: empirical standard error; CP (%): coverage probability of the 95% confidence interval.

| $\eta$ | Method | Bias | SE | CP |
|---|---|---|---|---|
| 4.33 | IVW | -87.8 | 0.040 | 0.0 |
| | MR-Egger | -79.9 | 0.065 | 0.0 |
| | MR-Median | -81.6 | 0.058 | 0.0 |
| | MR-RAPS | 32.7 | 17.548 | 96.6 |
| | dIVW | -181.3 | 93.937 | 96.7 |
| | pIVW | -1.7 | 0.392 | 94.4 |
| 9.52 | IVW | -76.8 | 0.038 | 0.0 |
| | MR-Egger | -63.2 | 0.059 | 0.1 |
| | MR-Median | -65.5 | 0.056 | 0.0 |
| | MR-RAPS | 3.6 | 0.194 | 95.8 |
| | dIVW | 3.5 | 0.194 | 95.9 |
| | pIVW | 0.3 | 0.185 | 94.7 |
| 21.85 | IVW | -59.2 | 0.034 | 0.0 |
| | MR-Egger | -42.1 | 0.052 | 1.4 |
| | MR-Median | -46.8 | 0.050 | 0.2 |
| | MR-RAPS | 0.1 | 0.092 | 95.2 |
| | dIVW | 0.2 | 0.090 | 95.3 |
| | pIVW | -0.5 | 0.089 | 95.0 |

### Web Table 9

The simulation with individual-level data. Comparison of the pIVW estimator ($\lambda_{opt}$ = 1) with other competing MR methods. The true causal effect $\beta = 0.5$ and balanced horizontal pleiotropy exists ($\tau = 0.01$). The IV selection threshold $\delta = \sqrt{2\log p}$. The simulation is based on 10,000 replicates. Bias (%): bias divided by $\beta$; SE: empirical standard error; CP (%): coverage probability of the 95% confidence interval.

| $\eta_\delta$ | Method | Bias | SE | CP |
|---|---|---|---|---|
| 6.76 | IVW | -6.8 | 0.172 | 90.7 |
| | MR-Egger | -41.7 | 0.532 | 87.7 |
| | MR-Median | -11.0 | 0.199 | 88.4 |
| | MR-RAPS | 2.6 | 0.192 | 92.7 |
| | dIVW | 3.7 | 0.198 | 95.5 |
| | pIVW | 0.5 | 0.189 | 95.1 |
| 10.26 | IVW | -8.2 | 0.111 | 90.8 |
| | MR-Egger | -43.7 | 0.289 | 83.8 |
| | MR-Median | -12.6 | 0.136 | 87.2 |
| | MR-RAPS | 0.7 | 0.123 | 93.4 |
| | dIVW | 1.0 | 0.124 | 95.5 |
| | pIVW | -0.4 | 0.122 | 95.1 |
| 17.84 | IVW | -8.2 | 0.070 | 89.9 |
| | MR-Egger | -48.7 | 0.181 | 71.0 |
| | MR-Median | -12.7 | 0.088 | 82.1 |
| | MR-RAPS | 0.3 | 0.078 | 94.1 |
| | dIVW | 0.4 | 0.078 | 95.2 |
| | pIVW | -0.1 | 0.077 | 95.0 |

### Web Table 10

Description of the GWAS datasets used in this paper:

| Trait | Dataset | Source | Population | Sample size (case/control) | Trait Description |
|---|---|---|---|---|---|
| Type 2 diabetes | Selection | GERA | European | 7,624/54,223 | ICD-9: 250 |
| | Exposure | UK BioBank | European | 6,024/106,314 | Self-reported: diabetes, type 2 diabetes<br>ICD-10: E11 |
| Dyslipidemia | Selection | GERA | European | 33,024/28,823 | ICD-9: 272 |
| | Exposure | UK BioBank | European | 16,818/95,520 | Self-reported: high cholesterol<br>ICD-10: E78 |
| Hypertensive disease | Selection | GERA | European | 31,000/30,847 | ICD-9: 401, 402, 403, 404 |
| | Exposure | UK BioBank | European | 32,689/79,649 | Self-reported: essential hypertension, hypertension, gestational hypertension /preeclampsia<br>ICD-10: I10, I11, I12, I13 |
| Peripheral Vascular Disease | Selection | GERA | European | 4,708/57,139 | ICD-9: 415, 440, 453 |
| | Exposure | UK BioBank | European | 1,816/110,522 | Self-reported: peripheral vascular disease, leg claudication/intermittent claudication, arterial embolism, pulmonary embolism +/- dvt<br>ICD-10: I26, I70, I82 |
| BMI | Selection | Akiyama et al | Asian | 173,430 | Body mass index |
| | Exposure | UK BioBank (GWAS round 2) | European | 359,983 | Body mass index |
| COVID-19 infection | Outcome | COVID19 Host Genetics Initiative (GWAS round 5)<br><br>*The datasets excluding UK BioBank are used to avoid possible sample overlap | European, African, Admixed American, Middle Eastern, South Asian and East Asian | 42,557/1,424,707 | Reported SARS-CoV-2 infection |
| Hospitalized COVID-19 | | | | 11,829/1,725,210 | Moderate or severe COVID-19 (i.e., hospitalized due to symptoms associated with the infection) |
| Critically ill COVID-19 | | | | 5,870/1,155,203 | Very severe respiratory confirmed covid (i.e., required respiratory support in hospital or deceased due to COVID-19) |

### Web Table 11

The numbers of IVs and the estimated effective sample sizes of five obesity-related exposures (i.e., peripheral vascular disease (PVD), dyslipidemia, hypertensive disease (HD), type 2 diabetes (T2D) and BMI).

| Outcome | Exposure | No IV selection | | IV selection threshold $\delta = \sqrt{2\log p}$ | |
|---|---|---|---|---|---|
| | | $p$ | $\hat{\eta}$ | $\hat{p}_\delta$ | $\hat{\eta}_\delta$ |
| COVID-19 infection | PVD | 1799 | 2.19 | 60 | 5.63 |
| | Dyslipidemia | 2338 | 36.42 | 143 | 37.57 |
| | HD | 2311 | 26.61 | 165 | 11.28 |
| | T2D | 2331 | 15.25 | 148 | 14.72 |
| | BMI | 1902 | 217.33 | 375 | 37.08 |
| Hospitalized COVID-19 | PVD | 1768 | 1.68 | 60 | 3.98 |
| | Dyslipidemia | 2068 | 37.67 | 147 | 39.52 |
| | HD | 2075 | 28.85 | 164 | 12.04 |
| | T2D | 2091 | 14.17 | 140 | 14.81 |
| | BMI | 1887 | 218.93 | 374 | 37.36 |
| Critically ill COVID-19 | PVD | 1781 | 1.86 | 61 | 4.28 |
| | Dyslipidemia | 2082 | 40.57 | 153 | 39.30 |
| | HD | 2060 | 30.29 | 172 | 11.62 |
| | T2D | 2042 | 13.48 | 145 | 14.97 |
| | BMI | 1889 | 218.25 | 377 | 36.98 |

### Web Figures

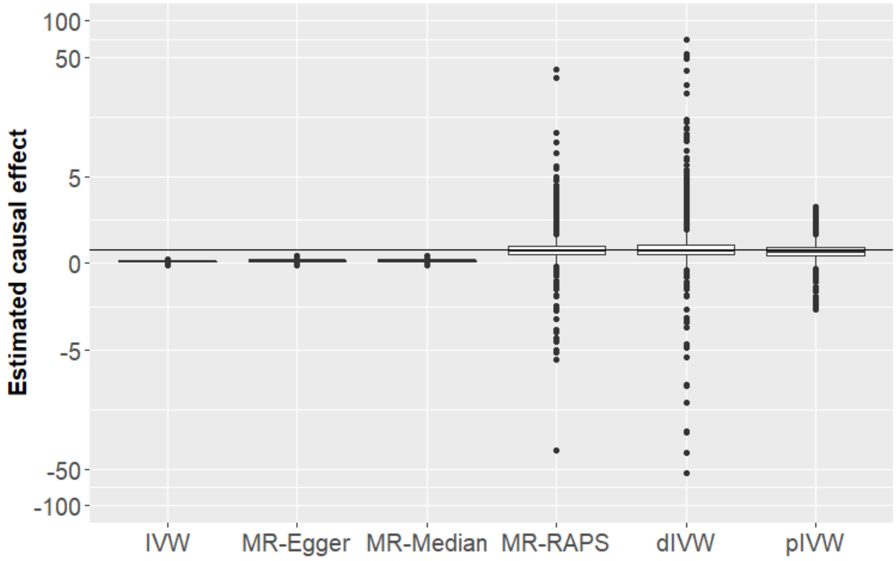

**Web Figure 1**. The box plot of the estimated causal effects of six methods. The true causal effect $\beta = 0.5$ (shown by the horizontal line). The effective sample size $\eta = 4.33$. No pleiotropy exists ($\tau = 0$). No IV selection is conducted. The simulation is based on 10,000 replicates. The pIVW estimator with the optimal $\lambda_{opt}$ = 1

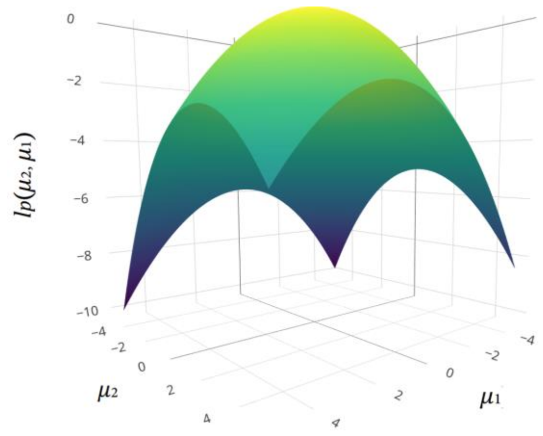

(a) $\lambda = 0$

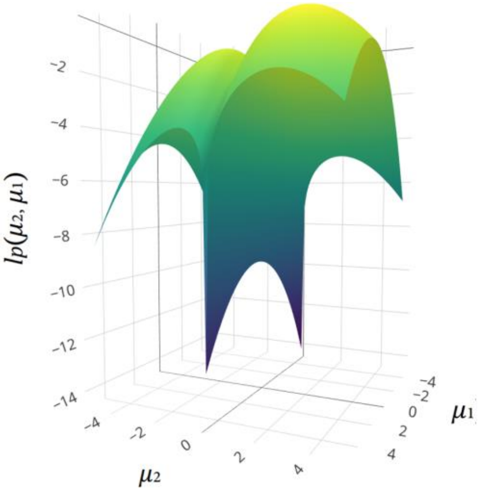

(b) $\lambda = 1$

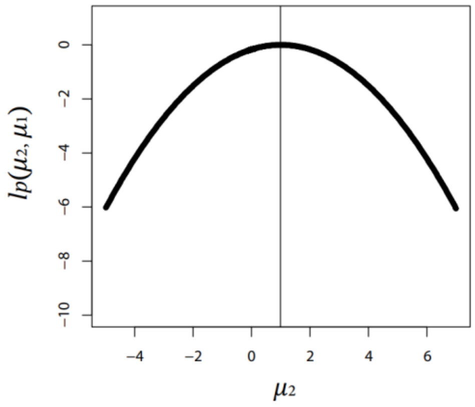

(c) $\lambda = 0$

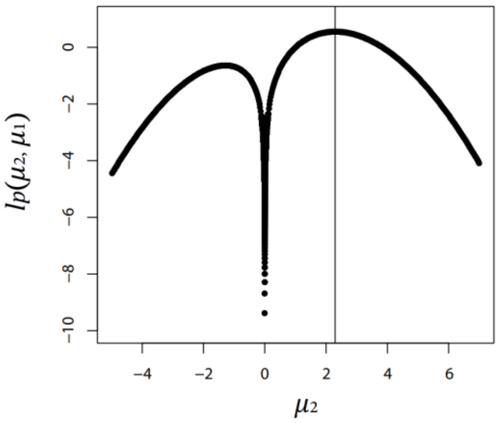

(d) $\lambda = 1$

**Web Figure 2.** The plots (a) and (b) show the penalized likelihood function $l_p(\mu_1, \mu_2)$ against $\mu_1$ and $\mu_2$ under $\lambda = 0$ and $\lambda = 1$, respectively, when $\hat{\mu}_1 = 0.5$, $\hat{\mu}_2 = 1$, $v_1 = 3$, $v_2 = 3$ and $v_{12} = 0.3$. The plots (c) and (d) shows $l_p(\mu_1, \mu_2)$ against $\mu_2$ with $\mu_1$ being fixed at the MLE estimates under $\lambda = 0$ and $\lambda = 1$, respectively. The vertical lines in (c) and (d) show the MLE estimates of $\mu_2$ under $\lambda = 0$ and $\lambda = 1$, respectively.

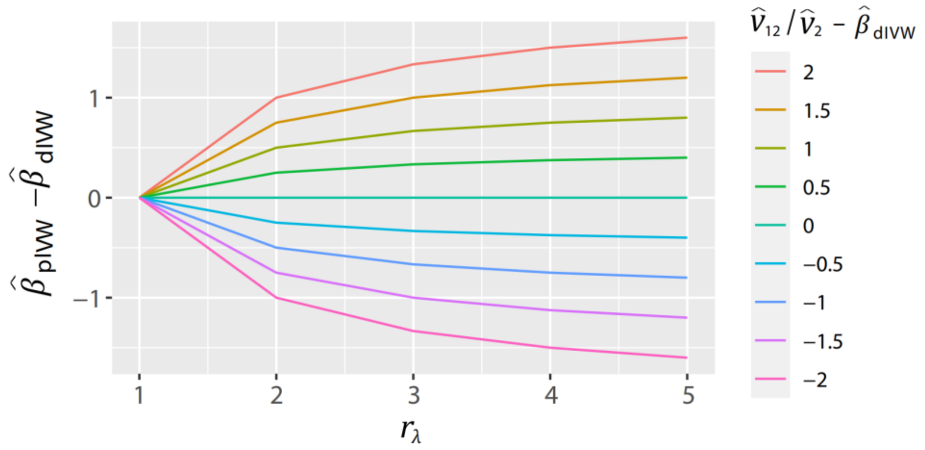

**Web Figure 3.** The plots of $\hat{\beta}_{\text{pIVW}} - \hat{\beta}_{\text{dIVW}}$ against $r_\lambda$ under different value of $\hat{v}_{12}/\hat{v}_2 - \hat{\beta}_{\text{dIVW}}$. When $r_\lambda = 1$, $\hat{\beta}_{\text{pIVW}} = \hat{\beta}_{\text{dIVW}}$. When $r_\lambda$ increases, the difference between $\hat{\beta}_{\text{pIVW}}$ and $\hat{\beta}_{\text{dIVW}}$ increases.

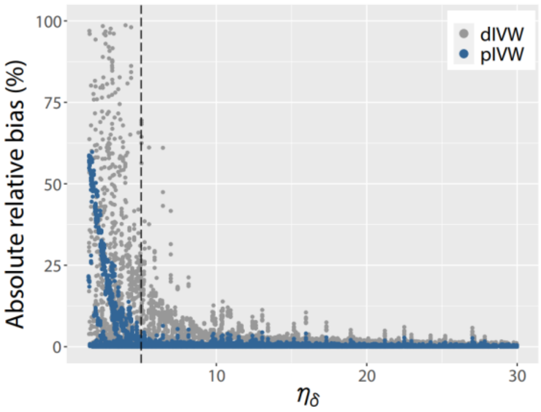

(a)

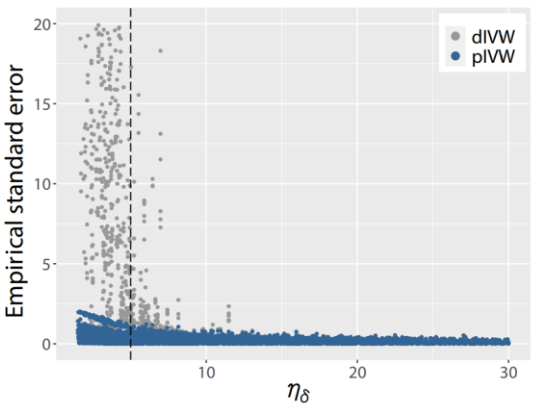

(b)

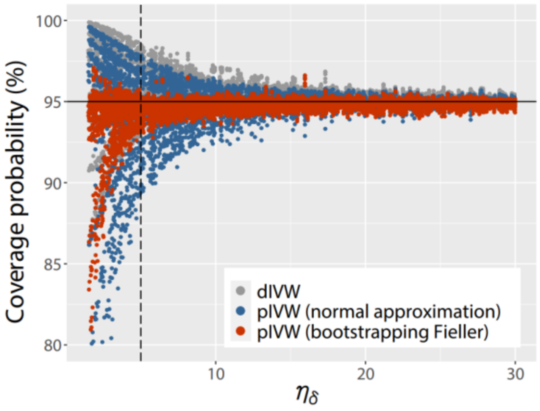

(c)

**Web Figure 4.** The plots of (a) absolute relative biases (%); (b) empirical standard errors; and (c) coverage probabilities (%) of the dIVW estimator and the pIVW estimator ($\lambda_{opt}$ = 1) against the effective sample size $\eta_\delta$. The dashed line shows $\eta_\delta = 5$. The dots represent the simulation results under different settings of parameters based on 10,000 replicates. There is no horizontal pleiotropy ($\tau = 0$). The IV selection threshold $\delta = \sqrt{2 \log p}$.

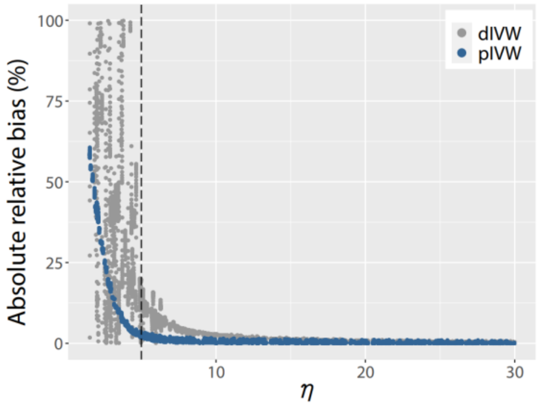

(a)

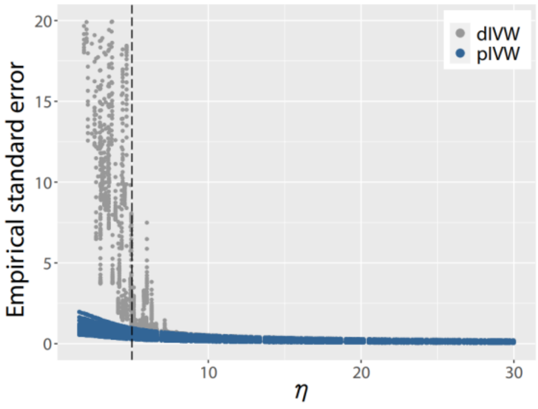

(b)

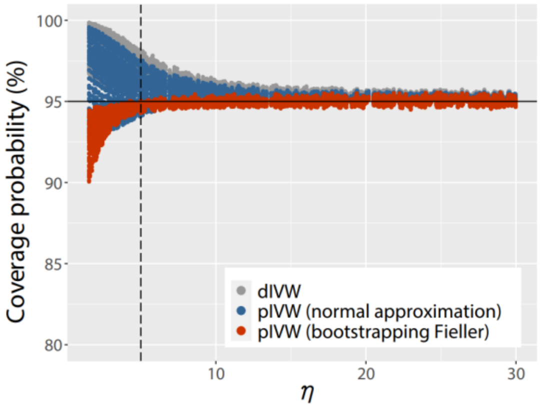

(c)

**Web Figure 5**. The plots of (a) absolute relative biases (%); (b) empirical standard errors; and (c) coverage probabilities (%) of the dIVW estimator and the pIVW estimator ($\lambda_{opt}$ = 1) against the effective sample size $\eta$. The dashed line shows $\eta = 5$. The dots represent the simulation results under different settings of parameters based on 10,000 replicates. The balanced horizontal pleiotropy has $\tau = 0.01$. There is no IV selection conducted.

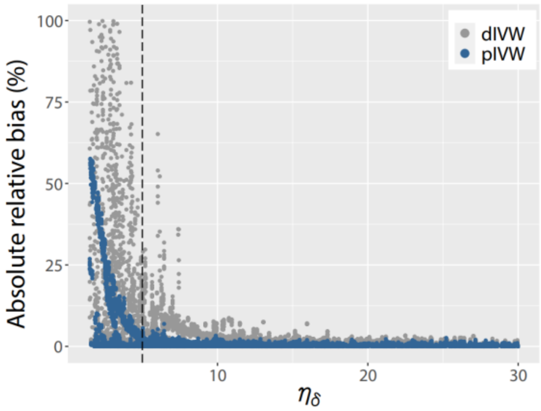

(a)

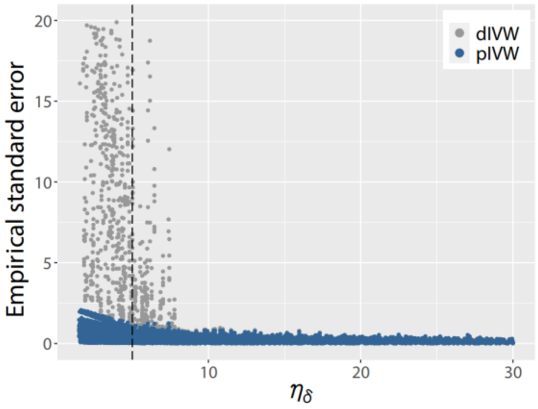

(b)

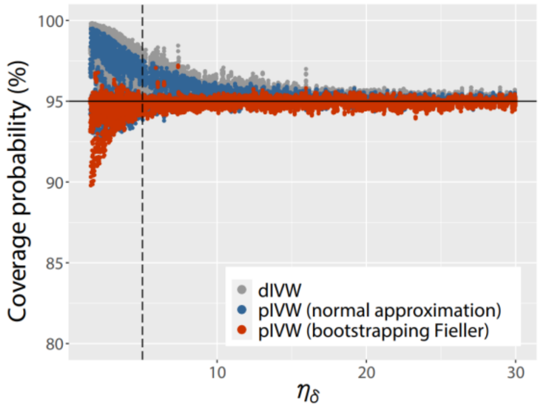

(c)

**Web Figure 6**. The plots of (a) absolute relative biases (%); (b) empirical standard errors; and (c) coverage probabilities (%) of the dIVW estimator and the pIVW estimator ($\lambda_{opt}$ = 1) against the effective sample size $\eta_\delta$. The dashed line shows $\eta_\delta = 5$. The dots represent the simulation results under different settings of parameters based on 10,000 replicates. The balanced horizontal pleiotropy has $\tau = 0.01$. The IV selection threshold $\delta = \sqrt{2\log p}$.

(a) Outcome: COVID-19 infection

| | IVW | MR-Egger | MR-Median | MR-RAPS | dIVW | pIVW |
|---|---|---|---|---|---|---|
| PVD | 0.003 (0.007) | 0.003 (0.01) | 0.005 (0.011) | 0.097 (0.223) | 0.102 (0.27) | 0.057 (0.14) |
| Dyslipidemia | 0.007 (0.013) | 0.010 (0.017) | -0.004 (0.025) | 0.017 (0.033) | 0.018 (0.032) | 0.018 (0.032) |
| HD | 0.025 (0.018) | 0.032 (0.025) | 0.019 (0.031) | 0.083 (0.055) | 0.074 (0.052) | 0.074 (0.052) |
| T2D | -0.005 (0.01) | -0.006 (0.013) | -0.012 (0.018) | -0.023 (0.042) | -0.023 (0.042) | -0.022 (0.041) |
| BMI | 0.095 (0.041) * | 0.119 (0.055) * | 0.038 (0.082) | 0.116 (0.05) * | 0.116 (0.05) * | 0.115 (0.05) * |

* P<0.05

Estimates: 0.08, 0.04, 0.00

(b) Outcome: hospitalized COVID-19

| | IVW | MR-Egger | MR-Median | MR-RAPS | dIVW | pIVW |
|---|---|---|---|---|---|---|
| PVD | 0.007 (0.013) | 0.022 (0.019) | 0.032 (0.02) | 0.379 (0.55) | 4.413 (95.942) | 0.219 (0.429) |
| Dyslipidemia | -0.005 (0.025) | -0.008 (0.033) | -0.013 (0.046) | -0.013 (0.063) | -0.011 (0.059) | -0.011 (0.059) |
| HD | 0.089 (0.033) * | 0.068 (0.047) | 0.109 (0.059) | 0.241 (0.095) * | 0.246 (0.093) * | 0.244 (0.093) * |
| T2D | -5e-04 (0.019) | -0.031 (0.026) | -0.038 (0.041) | -0.003 (0.085) | -0.002 (0.085) | -0.002 (0.083) |
| BMI | 0.382 (0.077) * | 0.455 (0.105) * | 0.549 (0.141) * | 0.466 (0.097) * | 0.468 (0.096) * | 0.468 (0.096) * |

* P<0.05

Estimates: 0.6, 0.4, 0.2, 0.0

(c) Outcome: critically ill COVID-19

| | IVW | MR-Egger | MR-Median | MR-RAPS | dIVW | pIVW |
|---|---|---|---|---|---|---|
| PVD | 0.009 (0.019) | 0.014 (0.029) | 0.030 (0.029) | 0.320 (0.578) | 0.464 (1.326) | 0.202 (0.449) |
| Dyslipidemia | -0.008 (0.036) | 0.014 (0.047) | -0.022 (0.066) | -0.020 (0.078) | -0.017 (0.077) | -0.017 (0.077) |
| HD | 0.092 (0.049) | 0.051 (0.07) | 0.081 (0.089) | 0.247 (0.133) | 0.244 (0.132) | 0.243 (0.131) |
| T2D | -0.033 (0.028) | -0.065 (0.041) | -0.042 (0.058) | -0.146 (0.124) | -0.139 (0.122) | -0.138 (0.12) |
| BMI | 0.366 (0.115) * | 0.431 (0.155) * | 0.391 (0.206) | 0.439 (0.141) * | 0.441 (0.14) * | 0.441 (0.14) * |

* P<0.05

Estimates: 0.4, 0.2, 0.0

**Web Figure 7.** Estimated causal effects and standard errors (in parentheses) of five obesity-related exposures (i.e., peripheral vascular disease (PVD), dyslipidemia, hypertensive disease (HD), type 2 diabetes (T2D) and BMI) on (a) COVID-19 infection, (b) hospitalized COVID-19, and (c) critically ill COVID-19. No IV selection is conducted. The pIVW estimator with the optimal $\lambda_{opt}$ = 1

(a) Outcome: COVID-19 infection

| | IVW | MR-Egger | MR-Median | MR-RAPS | dIVW | pIVW |
|---|---|---|---|---|---|---|
| PVD | 0.121 (0.034) * | 0.163 (0.043) * | 0.076 (0.065) | 0.278 (0.091) * | 0.297 (0.116) * | 0.272 (0.098) * |
| Dyslipidemia | 0.027 (0.025) | 0.023 (0.032) | -0.004 (0.03) | 0.031 (0.027) | 0.030 (0.027) | 0.030 (0.027) |
| HD | 0.013 (0.037) | 0.022 (0.051) | 0.039 (0.054) | 0.017 (0.046) | 0.016 (0.046) | 0.016 (0.045) |
| T2D | -0.010 (0.022) | -0.066 (0.029) * | -0.048 (0.035) | -0.012 (0.028) | -0.012 (0.029) | -0.012 (0.028) |
| BMI | 0.109 (0.056) | 0.123 (0.08) | 0.036 (0.095) | 0.117 (0.061) | 0.116 (0.06) | 0.116 (0.06) |

* P<0.05

Estimates: 0.2, 0.1, 0.0

(b) Outcome: hospitalized COVID-19

| | IVW | MR-Egger | MR-Median | MR-RAPS | dIVW | pIVW |
|---|---|---|---|---|---|---|
| PVD | 0.233 (0.055) * | 0.292 (0.071) * | 0.367 (0.098) * | 0.419 (0.217) | 0.670 (0.317) * | 0.594 (0.245) * |
| Dyslipidemia | -0.001 (0.038) | 4e-04 (0.05) | -0.013 (0.057) | -0.002 (0.042) | -0.002 (0.042) | -0.002 (0.042) |
| HD | 0.135 (0.065) * | 0.199 (0.091) * | 0.132 (0.099) | 0.161 (0.08) * | 0.163 (0.079) * | 0.163 (0.078) * |
| T2D | -0.026 (0.041) | -0.098 (0.054) | -0.047 (0.066) | -0.032 (0.052) | -0.033 (0.053) | -0.033 (0.052) |
| BMI | 0.371 (0.1) * | 0.563 (0.14) * | 0.361 (0.176) * | 0.397 (0.106) * | 0.397 (0.106) * | 0.397 (0.106) * |

* P<0.05

Estimates: 0.6, 0.4, 0.2, 0.0

(c) Outcome: critically ill COVID-19

| | IVW | MR-Egger | MR-Median | MR-RAPS | dIVW | pIVW |
|---|---|---|---|---|---|---|
| PVD | 0.228 (0.086) * | 0.291 (0.111) * | 0.381 (0.135) * | 0.555 (0.265) * | 0.613 (0.308) * | 0.549 (0.25) * |
| Dyslipidemia | 0.016 (0.056) | 0.015 (0.073) | -0.026 (0.076) | 0.016 (0.06) | 0.017 (0.062) | 0.017 (0.062) |
| HD | 0.108 (0.098) | 0.093 (0.136) | 0.087 (0.158) | 0.130 (0.122) | 0.133 (0.111) | 0.132 (0.111) |
| T2D | 0.025 (0.066) | -0.072 (0.088) | -0.036 (0.089) | 0.058 (0.092) | 0.032 (0.089) | 0.031 (0.088) |
| BMI | 0.292 (0.149) * | 0.450 (0.207) * | 0.379 (0.251) | 0.309 (0.158) | 0.311 (0.159) | 0.311 (0.159) |

* P<0.05

Estimates: 0.6, 0.4, 0.2, 0.0

**Web Figure 8**. Estimated causal effects and standard errors (in parentheses) of five obesity-related exposures (i.e., peripheral vascular disease (PVD), dyslipidemia, hypertensive disease (HD), type 2 diabetes (T2D) and BMI) on (a) COVID-19 infection, (b) hospitalized COVID-19, and (c) critically ill COVID-19. The IV selection is conducted at threshold $\delta = \sqrt{2 \log p}$. The pIVW estimator with the optimal $\lambda_{opt} = 1$
